# Quantifying physical degradation alongside recording and stimulation performance of 980 intracortical microelectrodes chronically implanted in three humans for 956-2246 days

**DOI:** 10.1101/2024.09.09.24313281

**Authors:** D. A. Bjånes, S. Kellis, R. Nickl, B. Baker, T. Aflalo, L. Bashford, S. Chivukula, M. S. Fifer, L. E. Osborn, B. Christie, B. A. Wester, P. A. Celnik, D. Kramer, K. Pejsa, N. E. Crone, W. S. Anderson, N. Pouratian, B. Lee, C. Y. Liu, F. Tenore, L. Rieth, R. A. Andersen

## Abstract

**Motivation:** The clinical success of brain-machine interfaces depends on overcoming both biological and material challenges to ensure a long-term stable connection for neural recording and stimulation. Therefore, there is a need to quantify any damage that microelectrodes sustain when they are chronically implanted in the human cortex.

**Methods:** Using scanning electron microscopy (SEM), we imaged 980 microelectrodes from Neuroport arrays chronically implanted in the cortex of three people with tetraplegia for 956-2246 days. We analyzed eleven multi-electrode arrays in total: eight arrays with platinum (Pt) electrode tips and three with sputtered iridium oxide tips (SIROF); one Pt array was left in sterile packaging, serving as a control. The arrays were implanted/explanted across three different clinical sites surgeries (Caltech/UCLA, Caltech/USC and APL/Johns Hopkins) in the anterior intraparietal area, Brodmann’s area 5, motor cortex, and somatosensory cortex.

Human experts rated the electron micrographs of electrodes with respect to five damage metrics: the loss of metal at the electrode tip, the amount of separation between the silicon shank and tip metal, tissue adherence or bio-material to the electrode, damage to the shank insulation and silicone shaft. These metrics were compared to functional outcomes (recording quality, noise, impedance and stimulation ability).

**Results:** Despite higher levels of physical degradation, SIROF electrodes were twice as likely to record neural activity than Pt electrodes (measured by SNR), at the time of explant. Additionally, 1 kHz impedance (measured in vivo prior to explant) significantly correlated with all physical damage metrics, recording, and stimulation performance for SIROF electrodes (but not Pt), suggesting a reliable measurement of *in vivo* degradation.

We observed a new degradation type, primarily occurring on stimulated electrodes (“pockmarked” vs “cracked”) electrodes; however, tip metalization damage was not significantly higher due to stimulation or amount of charge. Physical damage was centralized to specific regions of an array often with differences between outer and inner electrodes. This is consistent with degradation due to contact with the biologic milieu, influenced by variations in initial manufactured state. From our data, we hypothesize that erosion of the silicon shank often precedes damage to the tip metal, accelerating damage to the electrode / tissue interface.

**Conclusions:** These findings link quantitative measurements, such as impedance, to the physical condition of the microelectrodes and their capacity to record and stimulate. These data could lead to improved manufacturing or novel electrode designs to improve long-term performance of BMIs making them are vitally important as multi-year clinical trials of BMIs are becoming more common.

## 1. Introduction

Long-term recording and stimulation stability of microelectrode arrays is a fundamental requirement for the clinical viability of intracortical brain-machine interfaces (BMIs). Such BMI devices hold great promise for use in therapeutic and restoration applications by recording cortical neural activity [1]. These signals can be processed to decode an extraordinary amount of detailed information: motor planning and intent [2]–[10], high-level cognitive goals [11], [12], speech and language [13], [14], and dysregulated neural activity [15]. Furthermore, BMI’s can write information into cortical networks through electrical stimulation, creating novel somatosensory percepts [16]–[18], and visual stimuli [19], [20].

Longitudinal stability of recorded cortical neural activity has been investigated in both human and non-human primates, demonstrating that signals can be recorded from chronically implanted electrodes for up to four years [21], [22]–[24]. However, longitudinal BMI performance from intracortical microelectrodes (such as the Neuroport “Utah” array, Blackrock NeuroTech, Inc.) varies widely between participants, as recording quality deteriorates over time [23], [25]–[28]. Similar trends have been observed from Utah slanted electrode arrays during long-term peripheral nerve implants in human subjects [29] and animal models [30]. Biotic factors, such as inflammation and scar tissue encapsulation of the electrode sites, as well as abiotic degradation of device materials and mechanical failure from micro-movements of the electrodes relative to the brain, are all potential degradation mechanisms for implanted microelectrodes [28], [31]–[33].

Biological responses to implanted microelectrode arrays include glial activity and encapsulation of the implant in a glial sheath in response to the device being a foreign body [34], [35]. These tissue responses can adversely affect the performance of the electrodes. For instance, a case study of a participant with a Utah array implanted for seven months revealed tissue damage correlated with decreased recording performance [36]. Implanted electrodes can also be displaced by meningeal tissue downgrowth, in some cases contributing to nearly 30% of chronic device failures in non-human primates [37]–[39]. Increased implantation durations have been associated with greater meningeal downgrowth, increasing distances from function neurons [40], [41] and complicating the electrical pathway [38], [42], [43].

Material degradation presents another significant hurdle. Parylene-C, a common coating material for electrodes, can crack under physiological conditions [31], [44]–[46]. Encapsulation damage has been associated with decreased electrode impedance in previous studies, and can depend on the materials beneath the encapsulation coating [47]. All studies reporting longitudinal impedance of the Neuroport array have demonstrated substantial decreases in impedance asymptotically approaching values much lower than just after implantation [23], [27]. Additionally, silicon dissolution in the harsh *in vivo* environment continues to be a critical issue for implanted silicon-based devices [45], [47], [48]. The mechanisms and kinetics for silicon degradation in vivo has not been fully determined, and likely depend on multiple factors such as composition of the biological milieu, doping of the silicon, and fabrication processes.

The effects of electrical stimulation on electrode materials and surrounding tissues are crucial considerations for the long-term viability of BMIs. Aggressive electrical stimulation can erode sputtered iridium oxide films (SIROF) [49], another common coating material for electrodes, leading to a loss of functionality [50], though recent results suggest that SIROF optimization is improving the charge that can be delivered without degradation [51], [52]. Mechanical failure of electrode tips potentially due to stimulation-induced degradation is another common problem [53], [54], and also observed on some control electrodes.

To evaluate the lifetime and performance of neural implants, several techniques have been utilized to quantify failure modes of chronically implanted electrodes. Measuring encapsulation of chronically implanted arrays (∼1000 days for Pt and ∼200 days for SIROF), researchers evaluated images procured with scanning electron microscopy (SEM) [55]. These preliminary results showed the prevalence of grossly damaged electrode tips correlated positively with duration of implant, in line with other pre-clinical studies [32], [54]. Additionally, researchers noted irregular damage linked to stimulation, but did not describe a causal relationship. Accelerated aging techniques have been employed by Takmakov et al., using reactive oxygen species to simulate long-term degradation in vitro towards the establishment of known degradation mechanisms [56], [57]. Additionally, a large number of animal studies have sought to correlatively link electrode performance with measurable degradation qualities such as impedance and known foreign body reactions and biochemical responses [31], [58]–[61]. In short, the clinical viability of BMIs hinges on overcoming both biological and material challenges to provide a long-term stable interface for neural recording and stimulation. The objective of this study was therefore to assess potential damage to chronically-implanted intracortical microelectrodes and correlate the observed damage with measurable data collected throughout the implants’ lifespans.

## 2. Methods

### Terminology

Due to overlap of common terminology, we define some terms for the purposes of this paper. An *electrode* will be defined as a single shank on an array, comprised of a silicon shaft, insulation, and tip metal. An *electrode site* is the area with tip metallization exposed by removal of the Parylene-C insulation. *Shank* will refer to the silicon shaft within an electrode, which is 1.0 mm or 1.5 mm in length for FDA cleared arrays (K042384, K070272, K110010). An *array* will be the set of electrodes in a grid, typically either 10×10 or 6×10 (Figure 1C,D,F,G). A *channel* will refer to data collected from a single electrode on an array. An *assembly* will be comprised of a Neuroport pedestal connected to one or multiple arrays via a lead (wire bundle) of silicone encapsulated gold wires. Blackrock NeuroTech, Inc. (Salt Lake City, UT, USA) manufactured each of these arrays and assemblies. The company product documentation refers to an assembly of a single array and pedestal as a NeuroPort Array (Figure 1E), and an assembly with multiple connected arrays to a single pedestal as a MultiPort Array.

**Figure 1.**
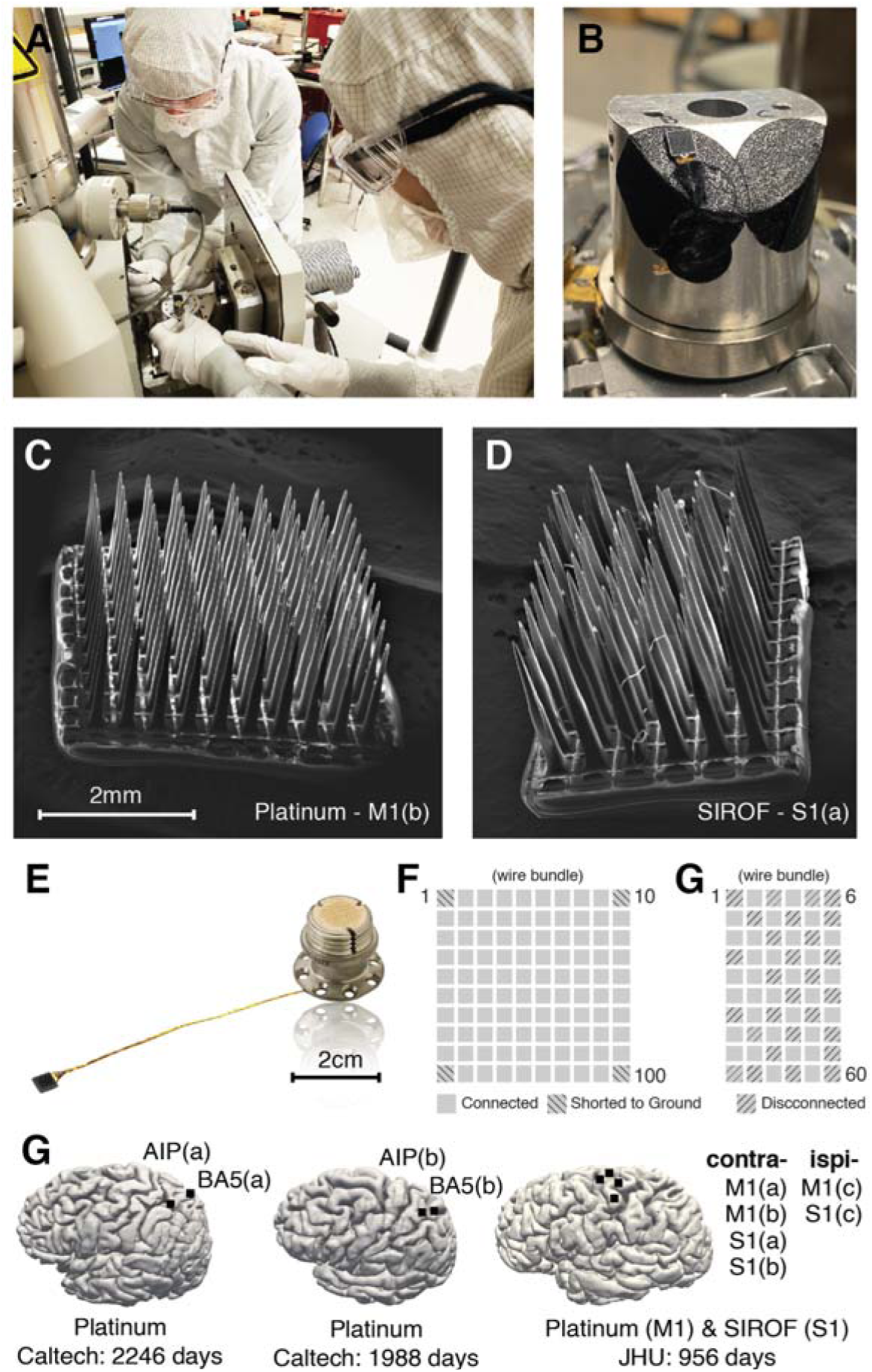
SEM images of devices and implant locations. Scanning electron microscopy was used to captured the physical damage of each array. **(A)** Scanning electron microscopy images were obtained at the Kavli Nanoscience Institute at Caltech and University of Utah NanoFab. **(B)** Each electrode was fixed to the “chuck” with carbon tape and wire bundles grounded. **(C)** Whole array image of platinum tipped array (M1b) from the JHU/APL participant, explanted after 2.6 years. **(D)** Whole array image of sputtered iridium oxide film (SIROF) tipped array (S1a) from the JHU/APL participant, explanted after 956 days. **(E)** NeuroPort image from the manufacturer, Blackrock Neurotech, Inc. Not shown, but two reference wires were attached to the pedestal for implantation near the array. **(F)** Channel layout of connected and disconnected electrodes for each type of array, and location of the wire bundle. **(G)** Implant locations for ten implanted arrays. Full surgical implant details and procedures can be found at their respective publications.

### Participants

Three human participants with C5-C6 level spinal cord injury were consented and chronically implanted with NeuroPort arrays [1], [42], [62]. All human subject research, experimental design and biomedical devices used for the participant in the Caltech/USC study were approved by the Institutional Review Boards (IRB) of the California Institute of Technology, University of Southern California, and Rancho Los Amigos National Rehabilitation Hospital for collection of this data in a registered clinical trial (NCT01849822). For the participant in the Caltech/UCLA study, all protocols were approved by Institutional Review Boards (IRB) of the California Institute of Technology and University of California Los Angeles (NCT01958086). For the JHU/APL participant, the study was conducted under an FDA Investigational Device Exemption; was approved by the FDA, the Johns Hopkins Institutional Review Board (JH IRB), and the NIWC Human Research Protection Office; and is a registered clinical trial (NCT03161067). All participants gave their written informed consent prior to participation in research-related activities.

### Devices

This work analyzed eleven microelectrode arrays as part of eight assemblies manufactured by Blackrock Neurotech for a total of 980 electrodes. Ten of these arrays were chronically implanted in human cortex for 956-2246 days, and one non-implanted array served as a control sample. Two tip metal types were analyzed: platinum (Pt) and sputtered iridium oxide film (SIROF). Four Pt arrays had a 10×10 grid layout, with 96 electrodes connected to a single pedestal (Figure 1F), and were implanted in human participants of the Caltech/USC [63] and Caltech/UCLA studies. Three *Multiport* assemblies were implanted in a human participant for the APL/JHU study, each comprised of one 10×10 Pt array (96 electrodes connected) and one 6×10 SIROF array (32 electrodes of the 60 connected) assembled to a single 128 channel Neuroport™ pedestal (Figure 1F,G) [18]. During the explant surgery, each array was disconnected from the pedestal by cutting the lead. One Pt array (10×10) was manufactured at a similar time as the Caltech implantations and remained non-implanted and in the original sterile packaging for a duration of 1337 days between manufacturer validation date and SEM analysis. This array was imaged to serve as a control sample. These assembles are laborious and time intensive to build, so the manufacturer validation date was used as the date of construction completion.

### Implantation

The participant at Caltech/USC had one Pt array in left anterior intraparietal area (AIP) and one Pt array in left Brodmann’s area 5 (BA5), which remained implanted for 2246 days. The participant at Caltech/UCLA had one Pt array in left AIP and one Pt array in left Brodmann’s area 5 (BA5), which remained implanted for 1988 days. One human participant was implanted at Johns Hopkins with three Multiport NeuroPort assemblies (one Pt array and one SIROF array each attached to a single pedestal) for a duration of 956 days. The participant had two Pt and two SIROF arrays in *left* primary motor and somatosensory cortex, respectively (dominant M1 and S1), and one Pt and one SIROF array in *right* M1 and S1, respectively. Full details of surgical methods and array locations (Figure 1G) can be found in prior articles for the first [63] and second Caltech participants [64], as well as the JHU/APL participant [18].

### Explant procedure

Teams at Caltech/USC, Caltech/UCLA and JHU/APL explanted the three groups of arrays. The precise handling of the explanted devices could not be determined in all circumstances, but either involved rinsing in water and storage, or sterilization with an enzymatic cleaner, rinsing and storage. If sterilized, explanted devices were soaked in Enzol™ (Advanced Sterilization Products, Irvine, CA, USA), an enzymatic detergent used to sterilize medical instruments. They were then rinsed thoroughly in pure water, air dried, and stored dry in closed containers. Prior work has carefully evaluated the effects of Enzol on the composition and chemistry of Utah arrays and Parylene films, and observed no changes [46]. Therefore, neither of the post-surgical handling procedures are anticipated to influence the results of this inspection. The lack of adhered soft tissue on the explanted devices suggests the enzymatic cleaning procedure is more likely for all arrays.

During the explant of the Caltech/USC participant (Figure 1: arrays AIP(a) and BA5), surgeons used a micro dissector to separate the wires from the cortex and arachnoid. They lifted each array circumferentially to extract each from the cortex. During the explant for the Caltech/UCLA study, both arrays were observed to be encapsulated with soft tissue. Removal of the devices left a clear impression in the tissue underneath, a common finding across all array locations (Figure S5).

Surgeons who performed the explant of the JHU/APL participant reported the skin was tightly adhered around the base of each pedestal, and indicated healthy skin tissue with no sign of infection. Arrays in the contralateral craniotomy (M1a,b, S1a,b) were extracted easily, with slight adherence to the cortical tissue. S1c was not removed intact, but M1c was extractable intact. Underlying cortical tissue appeared healthy with some fibrous tissue (Figure S5). Most electrode arrays had some scar encapsulation.

### SEM Characterization

Scanning electron microscopy images were obtained at the Kavli Nanoscience Institute at Caltech (FEI Quanta 200, Thermofisher, Hillsborogh, MI) and University of Utah NanoFab (FEI Quanta 600, Thermofisher, Hillsborogh, MI) (Figure 1A, Figure S4). Each of the 980 electrodes was imaged at 500x, 1200x, and 2500x with simultaneous acquisition of backscattered and secondary electron signals, resulting in nearly 6000 images. The images were primarily collected using a 20 kV primary beam energy in high-vacuum conditions using solid-state detectors on the pole-piece for backscattered and Everhardt-Thornley detectors for secondary images. Working distances of 10 to 20 mm were used to enable imaging of both the whole array at low magnification, and the individual electrodes at higher magnifications. Backscattered imaging provides significant Z contrast strongly facilitating discrimination of silicon, Pt or SIROF tip metallization, Parylene-C, and silicone materials. All images were collected without coating to improve surface conduction in order to maintain backscattered imaging contrast and to avoid alteration of the surface morphology. Occasional low-vacuum conditions were with the Large Field Detector (LFD) for secondary imaging when persistent charging disrupted imaging. Characterization of the arrays by these methods resulted in a dataset with nearly 6000 images. Each electrode was fixed to the “chuck” with carbon tape and wire bundles grounded to the best possible extent with carbon tape over the wirebundle and its cut-free end (Figure 1B).

### Longitudinal Neural Data Collection

Throughout the lifetime of each implanted and connected electrode, impedance data and broadband electrophysiological waveforms were recorded at regular intervals using the Blackrock Cerebus Neural Signal Processor (NSP) to assess the electrode functionality and performance (Figure 8). Electrodes at the four corners of all 10×10 Pt arrays were connected to patient ground, and 28 of 60 on the SIROF arrays were not electrically connected (Figure 1F,G). Thus, no electrophysiological data were recorded from these electrodes, but they were imaged by SEM.

The magnitude of impedance was measured using the impedance measurement function of the Blackrock Cerebus and patient cable. Measurements used a 1kHz sine wave with a magnitude of 10 nA peak-to-peak for 1 second, and the resulting voltage at electrodes recorded. The system cannot measure impedances <30 kΩ, and accuracy in the lower range of impedances deteriorates. This can impact the ability to accurately measure low impedance SIROF electrodes, particularly after long in-dwelling periods during which the impedances notably decrease.

Broadband electrophysiological waveforms were digitized at a 30 kHz sampling rate and hardware bandpass filtered between 0.3 - 15 kHz. A software bandpass filter (0.25 - 5 kHz) were applied to broadband signals to assess each electrodes’ ability to record individual action potentials from surrounding neurons. Baseline noise was longitudinally measured through the life of the implant as the estimated root-mean-square (RMS) of the software filtered signal (Figure 8D) [65], [66].

### End-of-study metrics for electrode functionality

Six experts systematically observed electron micrographs of the electrodes regarding five metrics of damage, scored as 1, 2, 3 or 4, from no damage (1) to severe damage (4) (Table 2, Figure 2A). Scores from all experts were averaged for each metric. *Metallization* quantified the loss of metal at the electrode tip (Figure S3). *Separation* assessed the amount of separation between the silicon shank and tip metal. Nominally, the metal should be in direct contact with the silicon shank as part of the device fabrication. *Insulation* assessed thinning, penetration, and/or cracking of the *poly(chloro-p-xylylene)* (Parylene-C) along the outside of the shank. Parylene-C is expected to be a smooth and conformal layer in the region imaged near the electrode site. *Shaft* assessed damage to the silicone shaft. *Growth* assessed tissue adherence or bio-material damage to the electrode.

**Table 1.** Timeline of electrodes.

| Array Type | Size | Study Location/Participant |  | Implant Location | Shank Length | Validation Date | Implant Duration | Stim? | SEM Imaging |
| --- | --- | --- | --- | --- | --- | --- | --- | --- | --- |
| Pt | 10x10 | Caltech/UCLA | P1 | AIP(a) | 1.0mm | 6/9/14 | 2246 days | no | 3/18/22 |
| Pt | 10x10 | Caltech/UCLA | P1 | BA5(a) | 1.5mm | 6/9/14 | 2246 days | no | 3/18/22 |
| Pt | 10x10 | Caltech/USC | P3 | AIP(b) | 1.5mm | 10/15/12 | 1988 days | no | 3/8/19 |
| Pt | 10x10 | Caltech/USC | P3 | BA5(b) | 1.5mm | 10/15/12 | 1988 days | no | 3/8/19 |
| Pt | 10x10 | Caltech | J1 | n/a | 1.5mm | 5/16/16 | 0 days (control sample) | no | 1/13/20 |
| Pt | 10x10 | JHU/APL | J1 | M1(a) | 1.5mm | 10/31/17 | 956 days | no | 3/18/22 |
| Pt | 10x10 | JHU/APL | J1 | M1(b) | 1.5mm | 10/31/17 | 956 days | no | 3/18/22 |
| Pt | 10x10 | JHU/APL | J1 | M1(c) | 1.5mm | 10/31/17 | 956 days | no | 3/18/22 |
| SIROF | 6x10 | JHU/APL | J1 | S1(a) | 1.5mm | 10/31/17 | 956 days | yes | 3/18/22 |
| SIROF | 6x10 | JHU/APL | J1 | S1(b) | 1.5mm | 10/31/17 | 956 days | yes | 3/18/22 |
| SIROF | 6x10 | JHU/APL | J1 | S1(c) | 1.5mm | 10/31/17 | 956 days | yes | 3/18/22 |

**Table 2.** Metric criterion for observed degradation.

| Metric Name | Score Range | Criterion for Intact (1) | Criterion for Degradation (4) |
| --- | --- | --- | --- |
| Metalization | 1 (intact) to 4 (severe degradation) | less than 10% tip metal loss | greater than 90% tip metal loss |
| Separation | 1 (intact) to 4 (severe degradation) | no visible separation | observed separation greater than 2 $\mu\text{m}$ |
| Insulation | 1 (intact) to 4 (severe degradation) | not observable or all deformations smaller than 5 $\mu\text{m}$ | any observed deformations greater than 15 $\mu\text{m}$ |
| Shaft | 1 (intact) to 4 (severe degradation) | tip was wholly intact | more than 12 $\mu\text{m}$ was fractured and/or missing |
| Growth | 1 (intact) to 4 (severe degradation) | no adhesions observed or all adhesions smaller than 5 $\mu\text{m}$ | any adhesion greater than 20 $\mu\text{m}$ |

**Figure 2.**
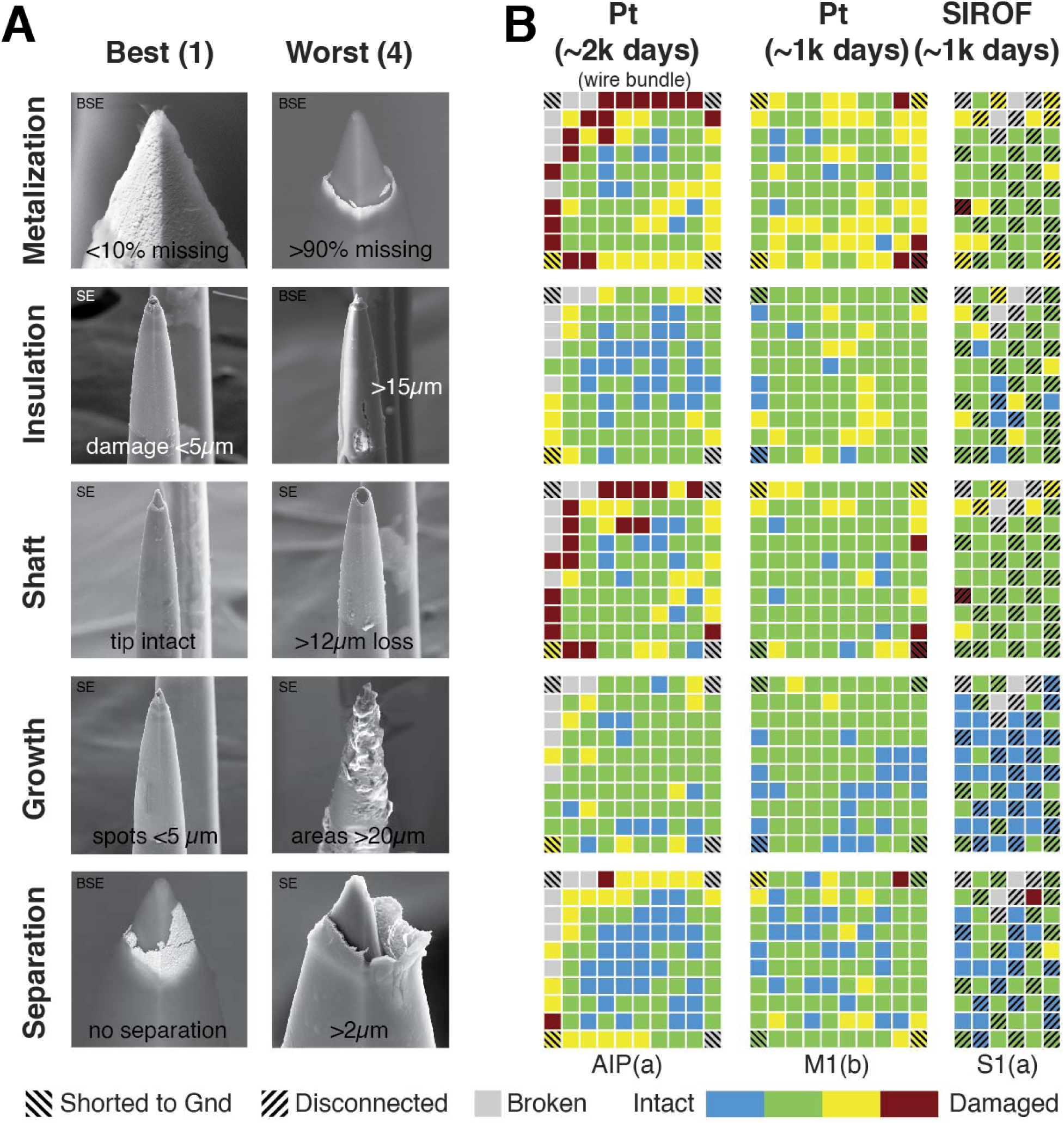
Electron microscopy to categorize electrode condition (end of study) Scanning electron microscope imagery was captured of each electrode at three focal zoom strengths (500x, 1250x, 2500x) with a standard image (SE) and backscatter (BSE). Backscatter images revealed metallization of the electrode tips while standard images captured all material. **(A)** Humans scored each electrode by visual inspection of the images on a scale from 1 (intact) to 4 (damaged) on five metrics: metallization, insulation, shaft, growth, and separation. **(B)** Example heatmaps scored by one reviewer for each damage metric; blue refers generally to an intact electrode, while red indicates damage. Disconnected electrodes are marked with diagonal lines. Shanks which were completely broken off were excluded from the analysis (marked in grey). The heatmaps correspond to a platinum array at 1988 days (array AIP(a)), a platinum array at 956 days (M1(b)), and a SIROF array at 956 days (S1(a)).

End-of-study impedance and neural data were measured and quantified prior to device explanation (Figure 3). For each electrode array implanted at Caltech, six datasets of broadband recorded electrical activity were collected in the two weeks prior to explant, with the same parameters as those collected longitudinally. Three datasets were collected during the two months prior to explant for the JHU/APL arrays. Impedances were collected alongside each of these datasets for all electrodes (Figure 3A). The recordings were used to determine noise and signal-to-noise ratio (SNR).

**Figure 3.**
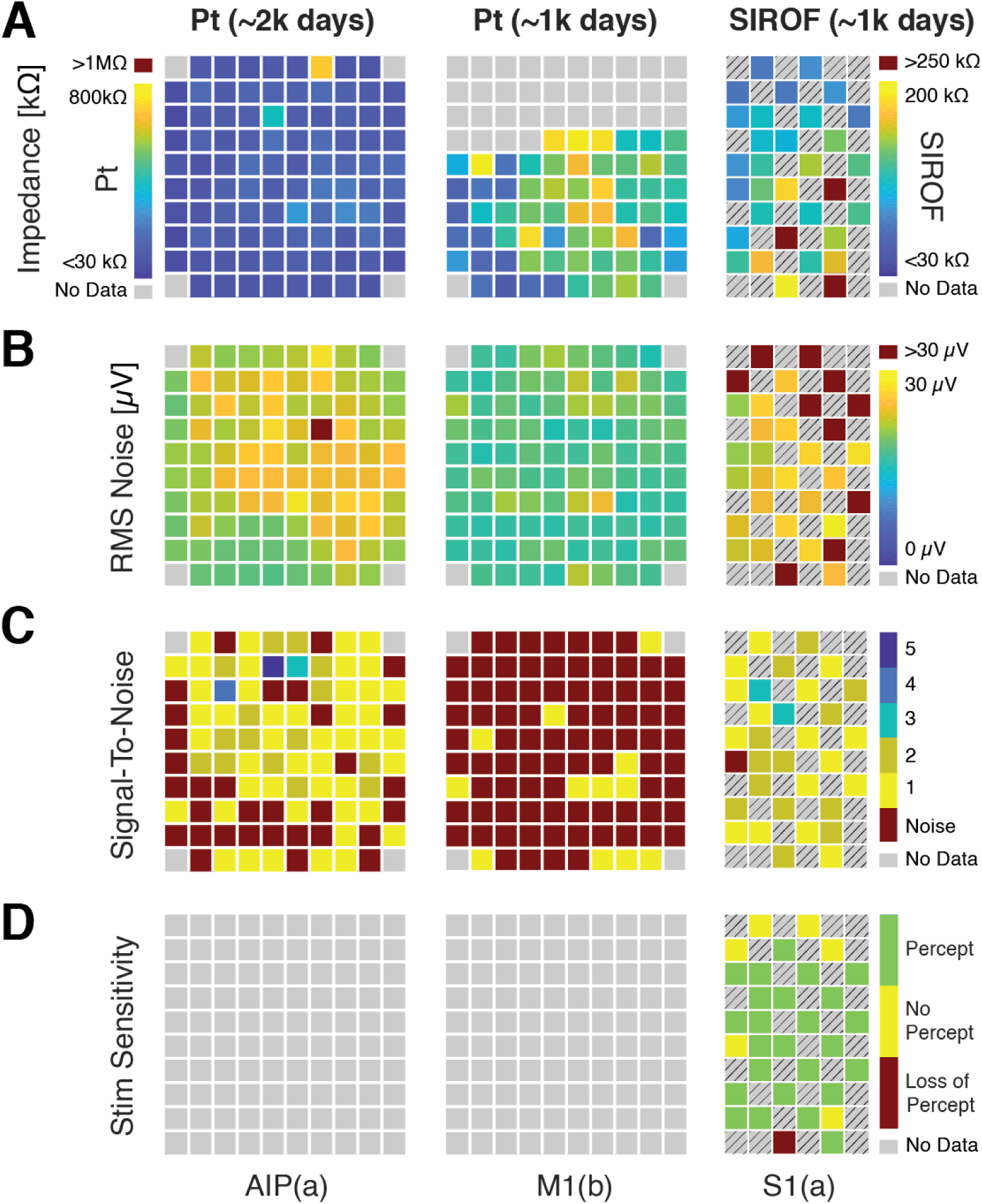
Functional assessment of electrode quality (at end of study) Prior to explant, several measurements were collected to assess functionality of each electrode’s condition: impedance, RMS noise, and signal-to-noise ratio. These data are presented from the electrode array AIP(a), M1(b), and S1(a), as identified in Figure 1. Disconnected electrodes are marked with diagonal lines and grey boxes are for electrode shanks with no data. **(A)** Example heatmaps of a Pt array at +5 years of implantation. Impedance is shown from 200 - 800 kΩ for the platinum arrays (AIP(a) and M1(b)) and from 30 - 250 kΩ for the SIROF array (S1(a)). **(B)** Example heatmaps of RMS noise for the example arrays. We plotted RMS noise from 0 - 30 μV. Greater than 30μV was shown in red. **(C)** Signal-to-noise ratio as a heatmap for the example arrays. SNR ranged from 0 – 5+. **(D)** We characterized each electrode as “able to generate a percept,” “never generated a percept,” or “generated a percept in the past, but unable to at end-of-life.” Platinum and disconnected electrodes were never stimulated, thus have no data.

Single and multi-unit neural action potentials were identified by extracting waveforms which negatively crossed a threshold of −3.5 times the RMS noise (Figure Figure 3B, 8A). From the average waveform of the largest single neuron identified on each electrode, the SNR values were calculated as the ratio of waveform’s mean peak-to-peak amplitude to its variance (Figure 3C, 8A).

The stimulation efficacy of the electrodes were evaluated by electrical stimulation patterns delivered through each of the connected SIROF electrodes. Using data collected during the last six months prior to explant, irrespective of stimulation pattern or frequency of use, each electrode was categorized. Firstly, if a sensation was able to be evoked up to the end of study; secondly, if a sensation was previously evocable, but unable at end-of-study; and thirdly, if no sensation had ever been evoked (Figure 3D). Total lifetime charge (milli-Coulombs [mC]) delivered through each electrode was also quantified to evaluate if the amount of stimulation correlated with electrode damage.

The electrodes could have been damaged at several different intervals during their lifetime. One potential point of damage could have occurred during surgical handling and insertion into tissue. Additionally, abiotic (device) degradation or mechanical failure during chronic in-dwelling could also contribute to changes in performance metrics. Thirdly, explant or post-explant handing could have fractured or damaged the electrodes. By quantifying impedance, SNR, and stimulation efficacy (stimulation arrays) recorded just prior to explant (Figure 3), we estimated the number of electrodes broken during the explant and/or transport phase (14/672 Pt) and (12/96 SIROF). Additionally, electrodes which were visibly destroyed (>50% of the shaft gone) were removed from the analysis (42/980 Pt and SIROF). Thus, this assessment of electrode damage focused on underlying abiotic mechanisms during the in-dwelling period, not surgical placement and removal techniques. Biotic degradation mechanism such as glial scar formation could not be analyzed, as this tissue was left intact during the explant procedure to minimize risks to study participants.

### Statistical analysis

We used Spearman’s correlation (with False Discovery Rate (FDR) correction for multiple comparisons) to test for significance on Figures 6 and 7. *Moran’s I* is a method of spatial autocorrelation [67], used to determine if damage indices are randomly arranged (complete spatial randomness (CSR) is the null hypothesis (*I = 0*)) or have a dispersed (*I = −1)* or correlated (*I = +1*) structure (Figure 6A). This represents the first time this spatial statistical technique has been applied to quantify the distribution of degradation of microelectrode arrays. For the violin plots in Figure 4, we used a non-parametric, two-tailed-sample bootstrapped comparison of overlapping confidence intervals to assess significance.

**Figure 4.**
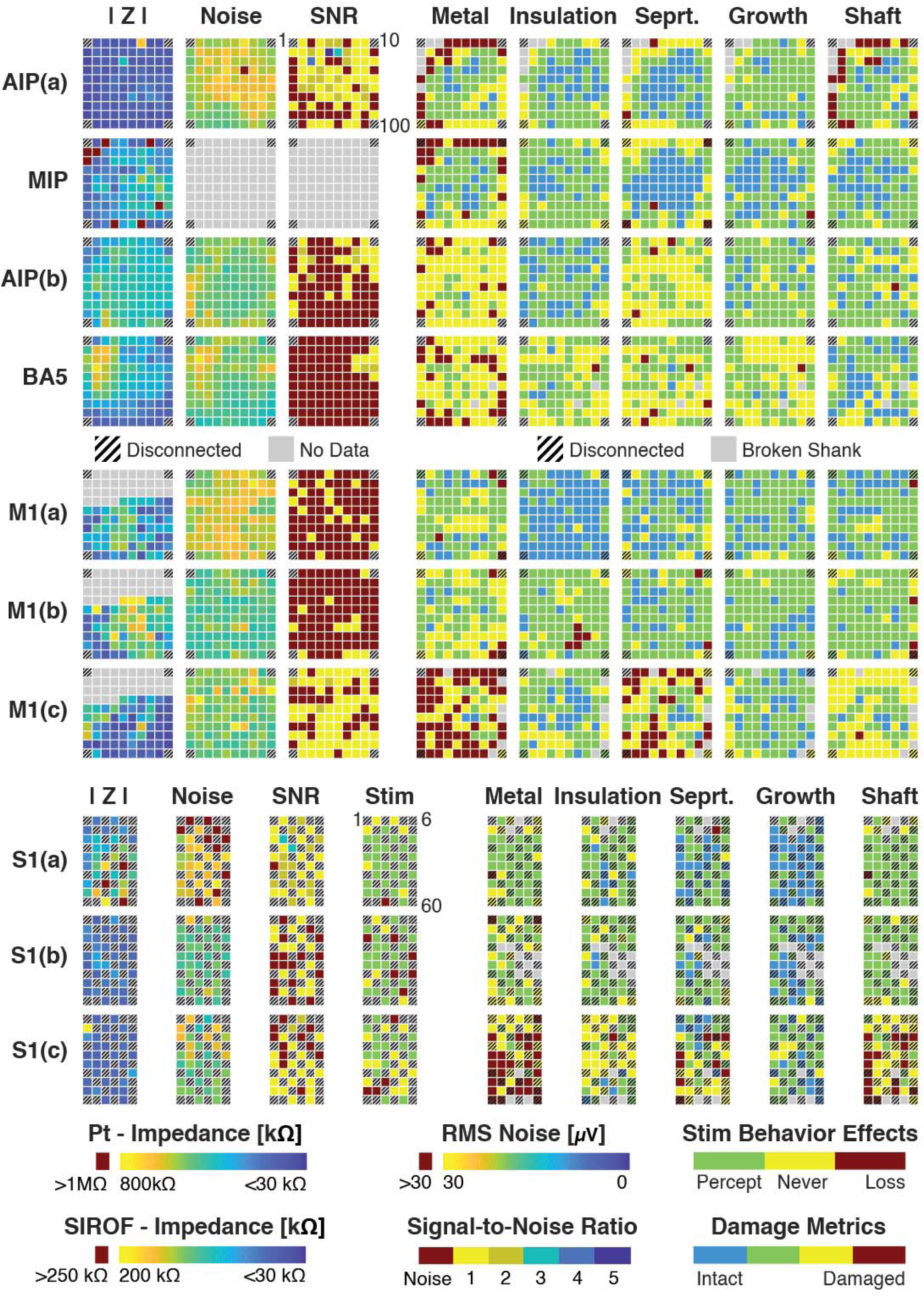
Heatmap of all electrodes and metrics (at end-of-study) Complete spatial maps for all arrays analyzed. Channel numbering starts from the top left corner, proceeding left to right, top to bottom. The wire bundle was connected to each array from the top of this layout. Scale bars indicating range of [kOhms] for impedance, μV for RMS noise, as well as average damage metrics, visual assessed from SEM images by experts. For all scale bars, blue refers generally to an intact electrode, while red indicates damage. Grey boxes have no data for Impedance (|Z|), RMS Noise, SNR, and Stim. Broken shanks are indicated with grey boxes for the visually-assessed damage metrics.

### Electrode groups

The 980 electrodes were grouped into five categories: platinum after ∼2000 days of implantation (n=389/400), platinum after ∼1000 days (n=393/400), stimulated SIROF (n=84/96), non-stimulated SIROF (n=72/84), and control platinum imaged 1342 days after manufacturer validation (n=100).

## 3. Results

This dataset from of eleven arrays provided a unique opportunity to compare degradation characteristics across several different metrics (implant duration, electrode tip material, electrical stimulation, and electrical connectivity), while chronically implanted in human cortex. For each array, we quantified the severity of observed damage of each metric for each electrode, scored by experts, and correlated it to recorded electrophysiological signals and behavioral data collected at the end of the study (Figure 4). Electrodes from the control array (not implanted) showed few signs of damage for any of the degradation metrics (Figure 5, purple).

**Figure 5.**
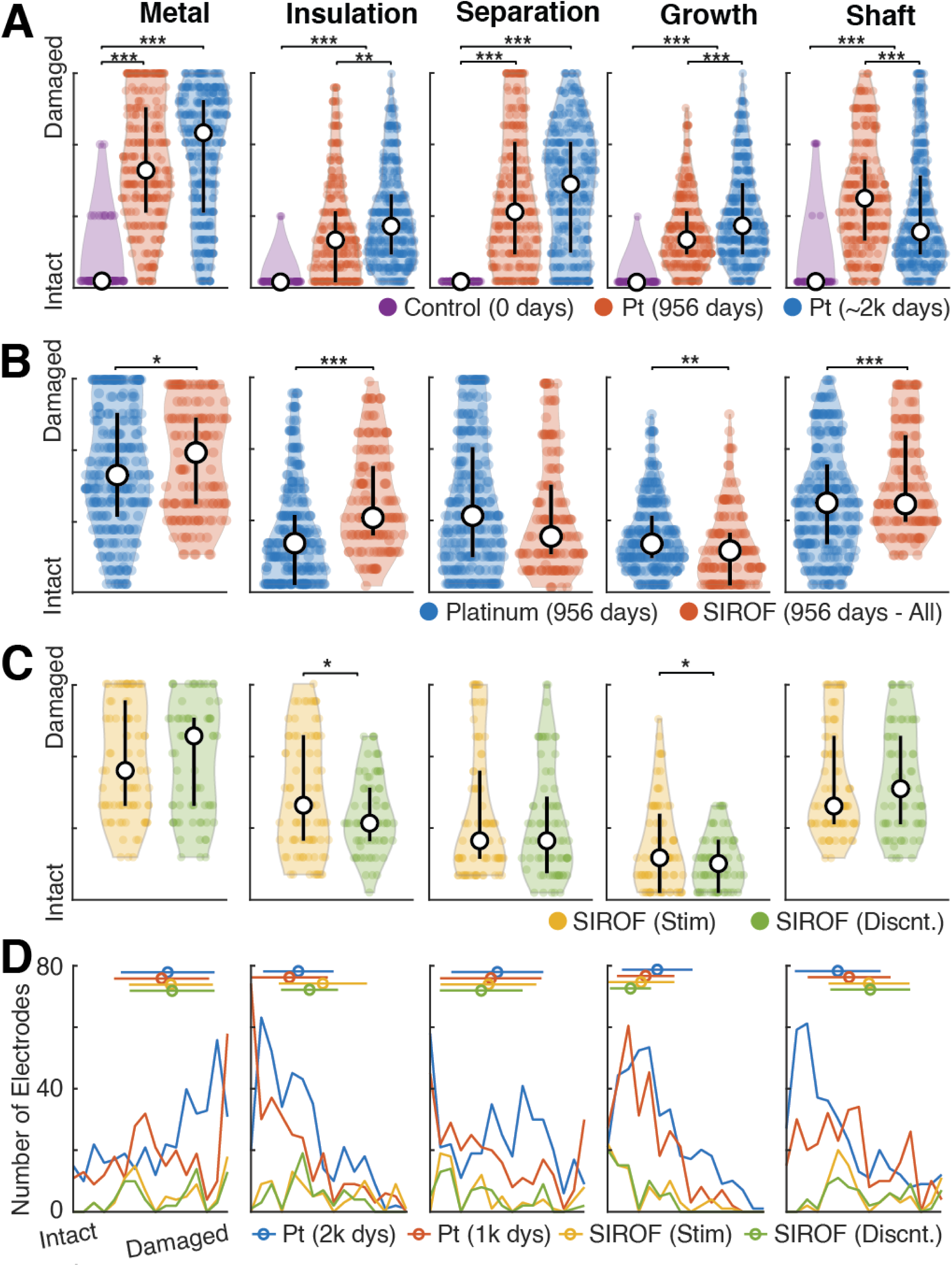
Degradation characteristics. Electrode scores for each metric visually scored by human experts were grouped according to implant duration (0 days: control, ∼1000 days, ∼2000 days), stimulation vs. disconnected, or tip material (Platinum vs. SIROF). Plots show the distribution of scores (violin plot), median score (white circle) and quartiles (black line). Significance was assessed via non-parametric two-sample bootstrapped confidence interval comparison and is indicated by black bars. Additional displays of array-specific information is provided in Figure S1. **(A)** Degradation on the platinum electrodes over time was observed for most metrics. Shaft damage was more prevalent on electrodes implanted for only ∼1000 days (JHU/APL participant) than those implanted for ∼2000 days (Caltech participants). **(B)** We observed significantly higher rates of damage to the metallization, insulation, and shaft on the SIROF shanks than then Pt shanks. Shaft damage was highest on the shanks implanted in the JHU/APL participant (all electrodes 956 days). **(C)** For SIROF electrodes implanted in the same participant for the same duration (2.6 yrs), we compared observed damage on electrodes through which stimulation was delivered and disconnected electrodes (see Figure 1G for spatial organization of SIROF electrodes). Two of the five damage metrics (insulation and growth) were statistically significantly worse for electrodes that had delivered stimulation vs. those that were discontinued. **(D)** Detailed histogram comparing all groups of electrodes. Mean and standard deviation are illustrated as a line and circle above the histogram. Further breakdown by array can be found in Figures S1 and S2.

### Implant duration

We observed an increase in damage correlated with implant duration (Figure 5A), through a comparison of three groups of platinum electrodes (0 days: control, ∼1000 days, ∼2000 days). As expected, a majority of the degradation metrics (metallization, insulation, separation, growth; Figure S1) showed a positive correlation. However, we also observed a significantly higher amount of shaft damage in the implanted Pt electrodes at ∼1000 days compared to ∼2000 days (Figure 4A – Shaft). The array M1(c) is observed to have notably higher levels of *shaft* damage serving as one mechanism for the differences. Small variation in shank thickness, likely due to variations in manufacturing, were observed in SEM images which is one possible cause for differences in shaft metrics.

### Platinum vs. SIROF

At the time of explant, SIROF electrodes were nearly twice as likely to record neural activity as Platinum electrodes (Figure 4, SNR), while implanted in the same participant for the same duration. For SIROF electrodes, 67/96 had an SNR > 1 vs. only 97/288 for Pt electrodes. Remarkably, we observed SIROF electrodes often had significant higher damage than their Pt counterparts (Figure 5B, p < 0.05, non-parametric, bootstrapped confidence intervals). Due to the heterogeneous types of distributions, these significance tests did not rely on an assumed distribution, however the mode/mean values did not always accurately represent the data (Figure 5D).

We observed SIROF electrodes had significantly more tip metal damage than Pt electrodes. However, there was no significant difference between metallization damage on stimulated SIROF electrodes compared to their disconnected counterparts (Figure 5C, Figure S2A). This may suggest stimulation was not the culprit for this increased metalization degradation between SIROF and Pt. In an additional analysis, we determined the amount of charge delivered also did not correlate with increased damage for any of our degradation metrics (Figure 7B, p > 0.05, Spearman’s rho). Damage to insulation and shaft were significantly higher (p < 0.05) for the SIROF electrodes (Figure S2A – Shaft, Insulation); however, tissue growth was significantly higher for the Pt electrodes. Additionally, Pt electrodes were more likely to have separation between the tip metal and shaft.

### Participant-specific effects

We also observed variability in the degradation metrics within participants for the same type of electrodes on different arrays (Figure 4, Metric: Metallization, Arrays: M1a, M1b vs. M1c). While the metallization, separation, and shaft damage for two of the Pt arrays (JHU/APL: M1a, M1b) were expectedly lower than the longer implanted electrodes (Figure S1B,C vs. S1D), the array implanted in the ipsilateral hemisphere (JHU/APL: M1c) was a clear outlier in damage, despite being the same electrode type (Pt) and having the same implantation duration (956 days) as M1a and M1b.

We also observed significant differences between platinum electrodes implanted for similar durations in different participants (Figure S1B,C). Metallization and separation damage were clearly higher for the arrays in the second Caltech participant (Figure S1C) than the first (Figure S1B).

### Stimulated vs. disconnected electrodes (SIROF)

Investigating the effects of stimulation in human cortex and the longevity of such electrodes was a key aim of this study. Of 60 electrodes on each SIROF array, there were 28 electrodes not electrically connected (floating) (Figure 1G). These electrodes provided a good control for potential damage caused solely by electrical connection, stimulation, or their combination. Due to the checkerboard pattern of connected and disconnected electrodes on the SIROF arrays (Figure 1G), stimulated and disconnected electrodes were evenly spatially organized.

Across the damage metrics from SEM inspection, we did not observe many significant changes in the degradation between stimulated and non-stimulated electrodes (Figure 5C, Figure S2). Slight but significant increases in insulation damage occurred, as well as an increase in observed growth on the stimulated electrodes.

### Type of damage correlated with stimulation

While inspection of the electrodes did not show significant differences in the *amount* of metallization damage between stimulated and disconnected electrodes (Figure 5C), we did identify significant differences in the *type* of damage (Figure 6B, Cracked vs. Pockmarked). The vast majority of the metallization damage observed on the platinum electrodes (Figure 6A) could be described as a “cracking” or “flaking” of the metal, peeling away from the silicon shaft (Figure 6D).

**Figure 6.**
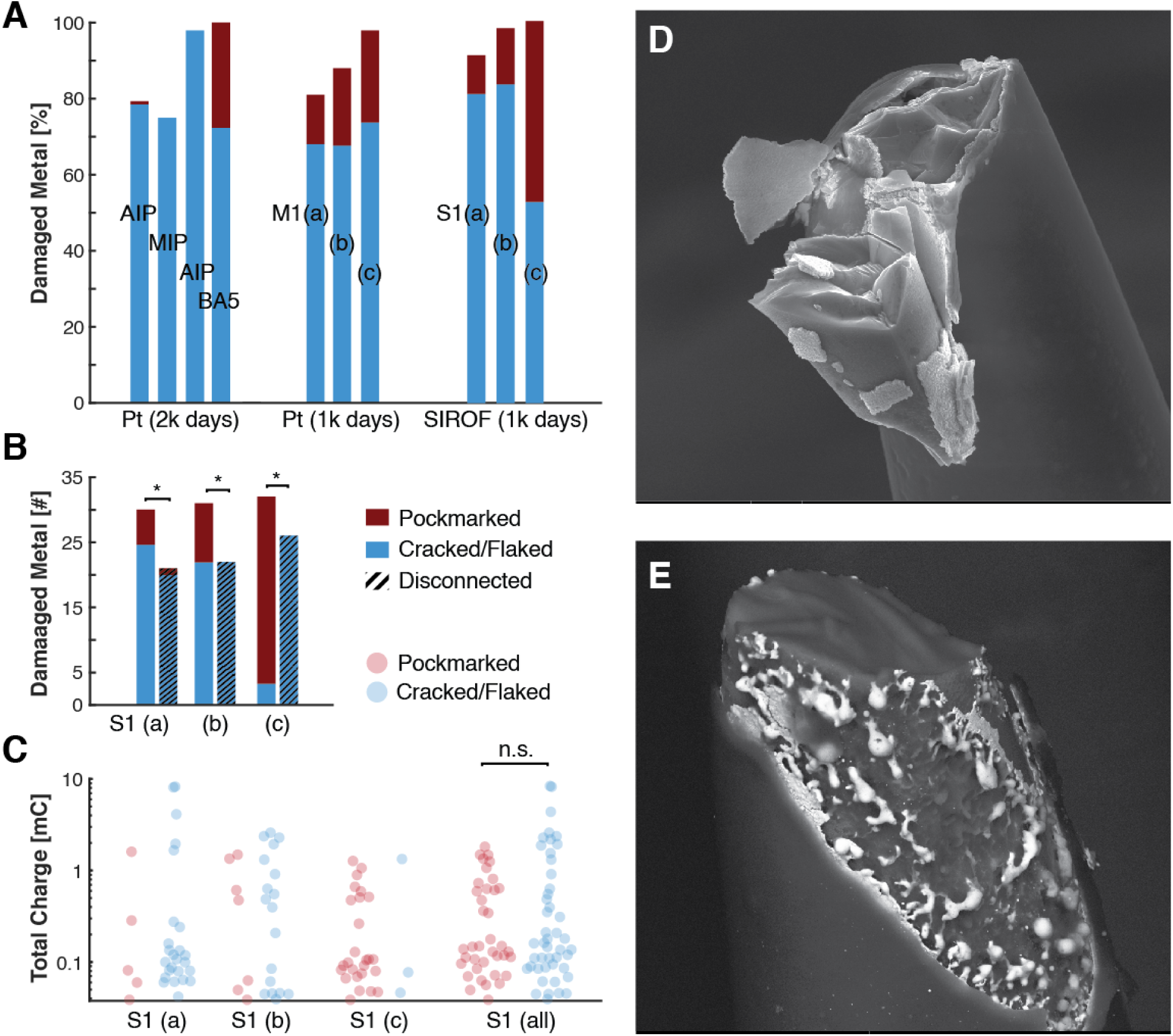
Tip metal damage. We observed and quantified two types of tip metal degradation: Cracking/Flaking (depicted in blue in the graphs) and Pockmarked (in red). **(A)** Pockmarked electrode tips were present on most arrays, but most prevalent on SIROF tips. Most arrays had more than 80% of electrodes with some metal damage. **(B)** When comparing electrodes which were stimulated to those which were disconnected, we can observe stimulation-specific effects. Pockmark degradation features were significantly more present on stimulation electrodes vs. disconnected ones. Stimulation electrodes were also more likely to have metal damage than the disconnected ones. **(C)** The total charge delivered through each damaged electrode in the SIROF arrays (S1(a), S1(b), S1(c), combined) is plotted in milliCoulombs (mC). The *type* of tip metal degradation was not correlated with the total amount of delivered charge. **(D)** SEM backscatter image of a SIROF electrode with cracked/flaking metal. See Figure S3 for more examples. **(E)** SEM backscatter image of a SIROF electrode showing pockmarked metal degradation.

We observed a different type of damage to the metallization, which we have named “pockmarked,” primarily occurring on the stimulated SIROF electrodes. This damage could also be described as “beading,” “melting,” or “congealing” of the tip metal. It is reminiscent of thermal damage, but there is not a commonly reported mechanism for such damage to occur within the in-vivo conditions that divides these two populations. No significant temperature changes occurred for this metal deformation. The potential across the electrochemical interface (E_mc_) is not expected to have been sufficient to drive, the metallurgical reactions that might result in deposition of metal, which is the mechanism by which rounded deposits of metal can form. Additionally, the damage resulted in exposure of the silicon shank under the metallization. It appears the SIROF material melted away and reattached itself to the silicon shaft in small nodules (Figure 6E), which speculatively could result from local gradients in the potential across the surface.

This observed “pockmarked” damage occurred with significantly higher frequency on stimulation electrodes vs. disconnected ones (Figure 6B), suggesting that stimulation may accelerate or enhance this form of metallization damage rather than the more typical “cracking” or “flaking.” However, it was also present in limited amounts on the platinum electrodes, thus stimulation is not the only way for this degradation mechanism to occur (Figure 6A). We also compared the amount of delivered charge through each stimulation electrode and did not see a significant between the metallization degradation types (Figure 6C), adding evidence that low current stimulation did not exacerbate damage beyond that caused by chronic implantation in human cortex.

### Spatially correlated damage

Most of the assessed damaged metrics were significantly spatially correlated (Figure 6A). Using Moran’s *I*, a measure of spatial auto-correlation (−1 indicates a fully dispersed pattern, 1 indicates full spatial correlation), we observed significant spatial correlations between SEM-inspected damage metrics and end-of-life measurements of electrode performance. The in-vivo milieu surrounding the electrodes is not expected to have large variations in composition that could lead to the correlation and grouping of damage observed. Such gradients could exist due to more chemical access to edge electrodes, and due to decreased of vascular access to the interior electrodes of the array. Edge electrodes are more likely to be damaged during fabrication and handling, and such damage could contribute to degradation in-vivo.

### Significant correlations between all damage metrics

Observed physiological damage of the electrodes affected multiple damage metrics rather than just a single feature (Figure 7C). For SIROF electrodes, the amount of growth on the electrode significantly correlated (rho > 0.5) with all other metrics, indicating a potential mechanistic driver for other degradation features. For both SIROF and Pt electrodes, metallization and separation had the highest correlation (rho > 0.75).

**Figure 7.**
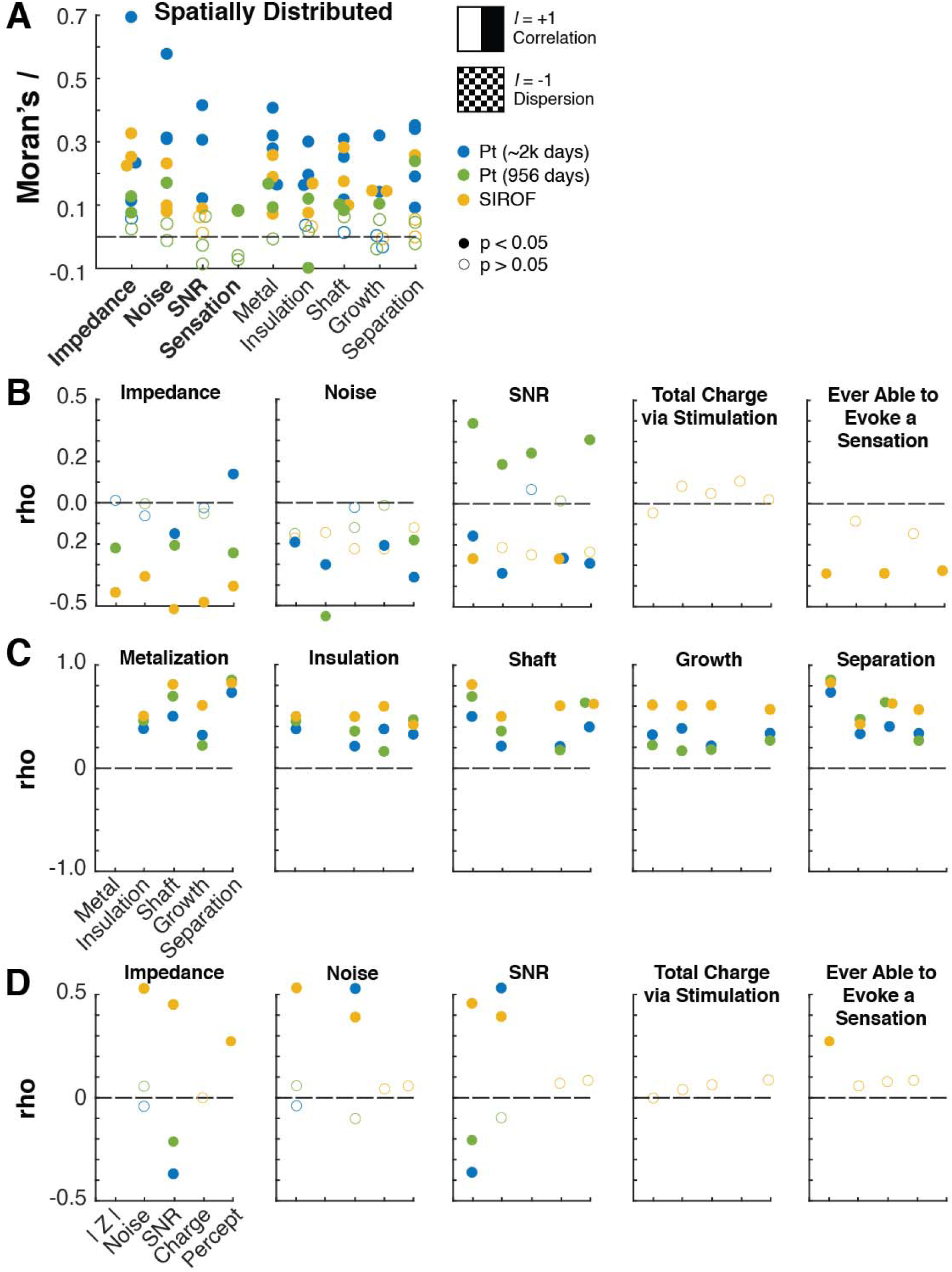
Analysis of observed damage. We evaluated relationships between damage metrics (metallization, insulation, shaft, growth, separation) and end-of-study metrics (SNR, impedance, noise). Filled in circles indicate statistical significance (p < 0.05). **(A)** Using Moran’s *I*, a measure of auto-spatial correlation, most metrics we observed were spatially correlated, suggesting damage was not due to implant/explant procedure but rather biological breakdown or tissue interaction. **(B)** We evaluated Spearman’s rho correlation between damage metrics and end-of-study metrics. Impedance significantly correlated with all damage metrics for SIROF electrodes, but was weakly correlated on Pt electrodes. The total charge delivered via stimulation did not correlate significantly with any of the damage metrics. **(C)** All damage metrics were positively correlated together, suggesting that no one metric occurred in isolation. Metal and separation metrics had the highest correlation for all groups (r > 0.75), while growth was positively correlated for SIROF electrodes for all other groups. This suggests a possible mechanistic driver for degradation for these types of electrodes. **(D)** SIROF electrodes had a significantly positive correlation between: impedance and RMS noise, Impedance and SNR, and RMS noise and SNR. Pt electrodes explanted at 956 days had a significantly negative correlation between Impedance and SNR. Pt electrodes explanted at 2246 days had a significantly negative correlation between Impedance and SNR, and a significantly positive correlation between RMS Noise and SNR. The total charge delivered via stimulation did not correlate with any of the other end-of-study measurements. Platinum and SIROF were oppositely correlated between impedance and SNR, the optimal impedance that correlates with detecting single units lays somewhere in the upper range of the SIROF electrodes and in the lower range for platinum, in a process that could involve tip materials and desinsulation length

### Correlation between electrode functionality and damage metrics

A primary aim of this study was to link observed electrode degradation to electrode functionality: recording neural activity and delivering stimulation (for SIROF only). We computed the correlation between each end-of-study electrophysiological measurement and each of the damage metrics from SEM (Figure 7B).

### Impedance correlates with SIROF electrode recording and stimulation performance and degradation

We found impedance to be highly correlated (Spearman’s rho, FDR correction) with all damage metrics observed by SEM for SIROF electrodes, but not for Pt electrodes (Figure 7B). This finding suggests impedance is a good measure of electrode damage for SIROF electrodes and should be continually measured throughout the implant lifetime to monitor electrode performance.

Impedance significantly correlated with recording performance for SIROF electrodes. Despite the higher noise of SIROF electrodes (Figure 8A, Noise), they achieved superior single unit recording characteristics (Figure 8A, Signal-to-Noise). Both noise and SNR significantly positively correlated with impedance measurements (Figure 7D, Impedance).

**Figure 8.**
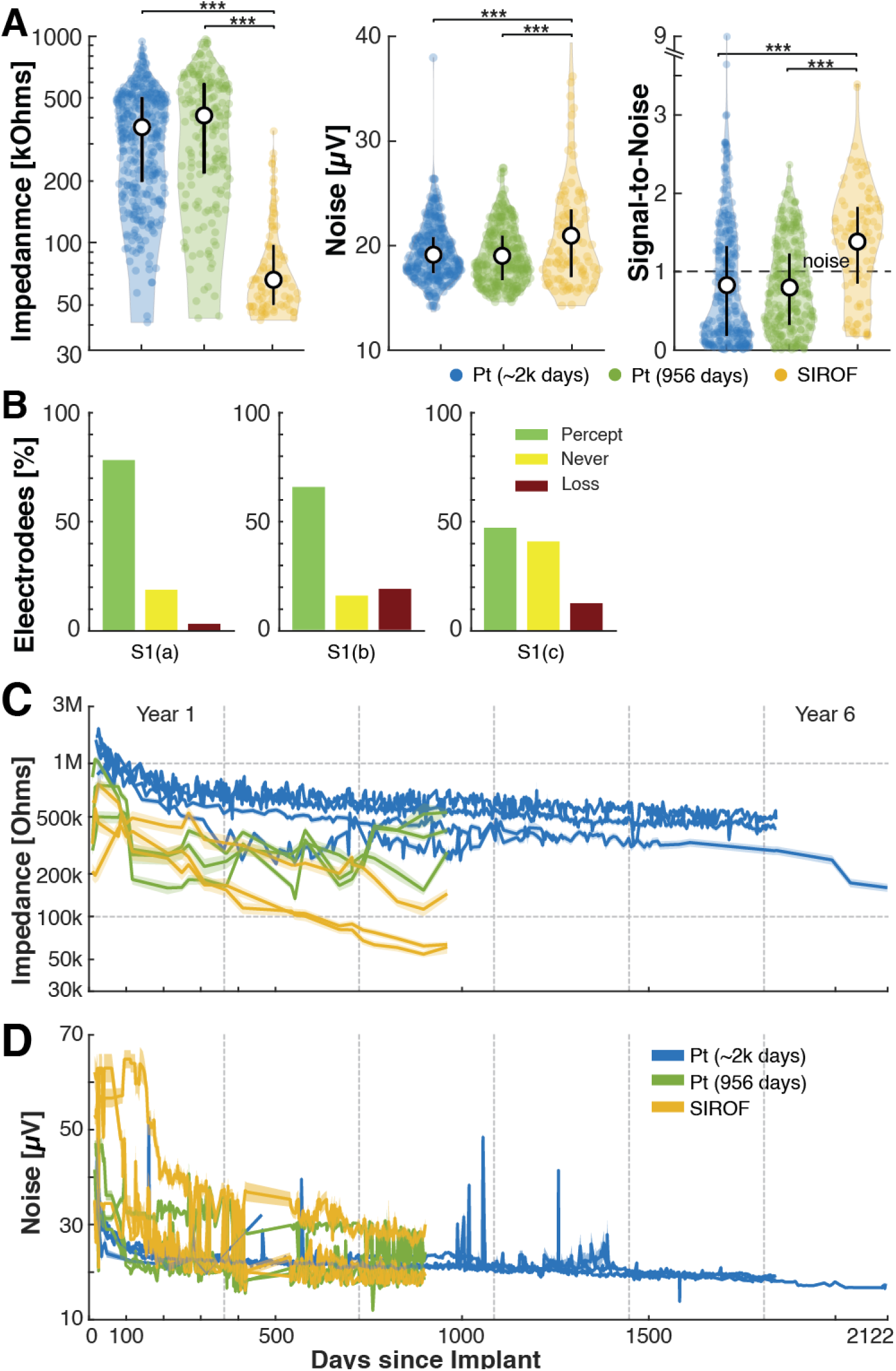
Electrode functionality. Longitudinal measures of electrode health were collected throughout the duration of the implantation and directly prior to explant. **(A)** As expected, measured impedance was significantly higher for both groups of Pt electrodes than SIROF at end-of-study. RMS noise was significantly higher for SIROF as well as recording performance, measured by the signal-to-noise. SNR was measured as the ratio of the mean peak-to-peak waveform to its variance. **(B)** For each of the SIROF arrays, the majority of the electrode were able to evoke a sensation at some point during its lifespan (green + red). A small number of electrodes stopped evoking sensations prior to explant (red). **(C)** Impedances for all electrodes were initially higher than their specified ranges immediately after implant. They quickly settled into their respective operating ranges within the 100 days. **(D)** Noise was assessed via an RMS calculation and remained relatively stable after the first 200 days post-implant. Values generally decreased with time, with occasional large outliers. These outliers often corresponded to noise in the signal, degradation of filament in the analog cables or other sources of environmental noise. Shaded error bars refer to 1 standard error of the mean of all electrodes on given array.

Impedance also significantly correlated with SIROF electrode stimulation performance (Figure 7D, Sensation). The functional ability of the SIROF electrodes to create a somatosensory percept (Figure 8B) was significantly negatively correlated to several damage metrics (Figure 7B), suggesting that increased electrode damage may decrease the likelihood of evoking a sensation. Importantly, we found that total charge delivered on stimulated SIROF electrodes did not significantly correlate any of the damage metrics (Figure 7B).

In contrast to the SIROF electrodes, impedance and SNR were negatively correlated for platinum electrodes (Figure 7D, Impedance); suggesting lower impedance Pt electrodes and higher impedance SIROF electrodes had the highest SNR values.

### Longitudinal electrode performance

All electrodes demonstrate a decrease in impedance and noise with time, consistent with analysis from other groups [21]. Such decreases would typically be attributed to encapsulation degradation, which could be consistent with decreased noise due to more averaging over a larger surface area. This suggests the optimal impedance and noise characteristics for detecting signal units, depend on a complex interplay between electrode material, exposed area, and associated noise characteristics.

Importantly, there are trends between longitudinally impedance and noise (Figure 8C,D). At early time points, the SIROF arrays have notably higher noise values and notably lower impedances. Most electrodes showed an initial high spike in impedance immediately post-implantation, with a gradual decay, reaching nominal ranges within the first year (200-800 kOhm for Pt, 80-200kOhms for SIROF). RMS noise generally settled into nominal ranges (14-28 µV) after the first 100 days (with the exception of array S1(c)). For the platinum arrays, this “settling” occurred much faster, generally within the first 25-50 days.

## 4. Discussion

A primary goal of this study was to quantify the condition and possible damage to chronically-implanted Neuroport electrodes and link observed damage to available, measurable data throughout the lifetime of the implant. We examined 980 electrodes implanted in three different BMI participants, comparing the effects of implant duration, electrode type, wired vs. unconnected, electrical stimulation, and the charge delivered by electrical stimulation. We imaged all electrodes using scanning electron microscopy and evaluated each for five components of damage, including metallization, insulation, shaft, growth and separation. We also captured end-of-study measurements just prior to explant: impedance, RMS noise, SNR, and stimulation functionality.

In general, we observed more severe damage on electrodes implanted for longer durations. Our data showed no correlation between electrode damage and the amount of delivered stimulation, but we did see a significant difference in the connected (wired) versus unconnected electrodes from the SIROF stimulation arrays. We also identified a new type of degradation feature (referred to as “pockmarked”), correlated with the prior delivery of stimulation, but also occurring on a few Pt electrodes. Impedance measurements were significantly correlated with 1) all five damage metrics observed by SEM for SIROF electrodes, 2) recording performance as measured by SNR, and 3) stimulation performance, suggesting impedance may be a valid performance measurement for these types of electrodes. Despite better recording performance at end-of-study (measured by higher SNR values), SIROF electrodes fared worse in damage compared to platinum electrodes; suggesting these electrodes may be better at capturing single unit activity, despite damage.

### Impedance strongly correlates with observed damage on SIROF electrodes but not Pt electrodes

In absence of any other defined metrics or observable variables, measuring electrode impedance has long been the gold standard in determining *in vivo* electrode health [31], [70]–[73]. However, as electrophysiologists very commonly note, correlation between the 1 kHz impedance magnitude and signal performance is tenuous at best [74], [75]. This study validates these fears with respect to platinum electrodes, for which impedance was weakly correlated with observed damage (Figure 7B – Impedance: blue, green). However, impedances collected from SIROF electrodes strongly correlated with all observed damage metrics (Figure 7B – Impedance: yellow). As expected from with previous studies, impedances generally decreased over time (Figure 8C), likely due to a combination of encapsulation [28], [75], metallization breakdown, increased exposed electrode surface area. This material and surface area exposed play an important role in such impedances changes, as has been described [47]. This longitudinal trend correlates with increased degradation over time [55], however in our electrode by electrode data, only impedance from SIROF electrodes reliably correlated with all damage metrics (Figure 7B).

### Proposed degradation mechanisms

We observed a variety of electrode metallization failure modes and sought to characterize a potential timeline and mechanisms for such degradation (Figure S3). We observed a high frequency of electrodes with highly damaged tip metal (Figures 3, Metallization); however, some electrodes remained in near perfect condition (Figure S3A). The electrodes ranked “Intact” (Figure S3A,B,C) exhibited no imperfections or slight damage, including the very top of the electrode and/or hairline fractures or holes in the metallization at the base of the tip (Figure S3C). While damage might be present below the metallization as has been observed previous [46], these results are suggestive that achieving improved stability for electrodes is possible. Whenever we observed the holes in the tip metal, we also observed etching of the silicon shank underneath, likely causing separation between the metal and the silicon (Figure S3D-I). With severe etching/erosion, we observed flaking of the tip metal, often with such remnant flakes observed in the images (Figure S3G-I). At the worst case, we observed nearly all the metal had been mechanically removed due to undercutting (Figure S3J) and/or the silicon shank had been severely etched (Figure S3K,L). In several cases, there appears to be consistently oriented fracture surfaces in the silicon shank of multiple electrodes potentially leaving them more susceptible to etching, contributing to large parts of the electrodes peeling away. Some evaluation of the timeline for such fractures can be achieved by careful evaluation of the fracture surfaces. Brittle fracture surfaces of exhibit mirror, mist, and hackle zones [76], with the latter having sharply defined features. Prior and current observations have shown silicon serves with smoothened features consistent with being etched in-vivo. These features are often coincident with separations between the metallization and silicon providing evidence that silicon dissolution contributes to smoothening features. Electrodes with both sharp and smoothened characteristics were observed, but quantifying such characteristics is beyond the scope of this work.

Some component of the observed variability of electrode longevity may be due to manufacturing differences (these would be an uncontrolled noise variable) [31]; however determination of this is beyond the scope of this dataset. Additionally, rigorous testing of our hypothesized mechanism of silicone corrosion would require significant experimentation best accomplished with bench studies such as electrochemical corrosion studies [77] to identify mechanism. The can be followed by animal studies to verify degradation mechanisms or mitigation strategies similar to prior studies with Utah arrays [46] and Michigan probes [48]. Factors such as the doping concentration, integrity of the tip metallization and encapsulation layers or potential other process factors might affect the longevity and performance of silicon based neural electrodes.

### Highly damaged electrodes can still yield good neural recordings

Some electrodes observed to have “high damage” captured quality neural signals during the end-of-study measurements. An example electrode (Figure S3 – Array AIP(a), Channel #5), scored the worst rating on metallization and shaft damage, and the second-worst rating on separation damage. However, during the end-of-study measurements, this electrode had an SNR of 2, meaning a significantly large single-unit waveform was recorded while impedance measurements appeared nominal.

Two justifications may align this incongruency. Firstly, physical characterization of electrical connectivity between metal and silicone may be difficult to accurately judge. SEM images obtained approximately 180-270 degrees of coverage of the round electrodes, leaving a small blind spot on the back side of the electrode preventing observations of regions potential conducive to recordings. A precise test of electrical functionality (i.e., impedance measurements post-implant) may more accurately capture such information; however, due to surgical requirements during explant, the wired connection between the pedestal and array is severed, making such measurements impossible. Secondly, small aspects of metal physically and electrically attached to the silicone shaft, but undercut surfaces and other features yield sufficient surface area to enable recordings. These results suggest only quite small electrode areas might be necessary to capture neural signals. In this best-case scenario, electrodes may be able to tolerate significant damage and yet remain viable for recording.

Whichever hypothesis future studies confirm, our observations do provide some reasoning behind the ability of electrodes to continue to function well into the realm of >1000 days of chronic implantation, despite known degradation [21]–[24].

### Limited degradation on stimulating electrodes

Across three stimulation arrays and five degradation metrics observed from SEM imaging, we found partial evidence of additional damage caused by stimulation (Figure 5, Figure S2). Electrodes which were stimulated, displayed significantly higher insulation damage and tissue growth adhered to the electrode. In contrast to previous findings from a much shorter time scale (182 days) [55], we did not observe an increase in metallization damage. Given the large changes in impedance and inflammation response during the first six months of implant, it is possible the electrode interface had not yet stabilized in the prior study [21].

While the frequency of metallization damage was not significantly different from stimulation vs. non-stimulation SIROF electrodes, our imaging has revealed a new type of electrode tip metal degradation, predominately observed on stimulating electrodes (Figure 6B). This “pockmarked” degradation is markedly different in visual characteristics to traditionally observed metal degradation which often appears as “cracked/flaking” (Figure 5D vs. 5E). This new “pockmarked” metallization damage was also observed on non-stimulated platinum and SIROF electrodes (Figure 5A,B), albeit at a much lower frequency of occurrence. We hypothesize stimulation may accelerate this damage type but not be the sole cause, since it is not unique to stimulated electrodes.

Furthermore, the total charge delivered through a stimulating (SIROF) electrode did not significantly correlate with any of our visually-assessed damage metrics (Figure 6B – Stimulation) or damage type (Figure 5C). Functional assessments of SIROF electrode performance (ability to evoke a sensation) also did not correlate with any end-of-study measurements or damage metrics, contrasting with some prior studies [78].

Some explant studies showed significant correlation between electrode metalization damage and stimulation [55]; despite the maximum total charge delivered was only 0.08 mC and implant duration was only 182 days. In our study, electrodes were stimulated up to 8 mC in total and implanted for 956 days, and they showed no such correlation on any array (Figure 6).

### Explant procedures

One limitation of the comparisons across the three participants is the differences in explant procedures. Given the three different clinical trial locations, the three different neurosurgeons involved, and the novelty of each participant’s surgical procedures, there existed no protocol from the manufacturer for removal and handling or storage of the arrays. While beyond the scope of this study, it may be beneficial for the manufacturer and other interested parties to develop a standardized explant procedure for device removal, treatment, storage, and end-of-study measurements to assess electrode degradation which may have occurred while implanted.

### Histology

Unlike other analyses of implantations occurring in to-be recessed tissue, the tissue in which these arrays were implanted was not available for resection or further analysis since the participants were still living at the time of explant. Previous analysis from our group on tissue damage from chronically implanted electrodes in human cortex showed increases in lymphocytic infiltrates, astrogliosis, and foreign body reaction at the implant site, as well as vascular disruption and signs of microhemorrhage from the implant procedure [36]. In all explant notations, the corresponding neurosurgeons observed the typical impressions left in the cortex after explant (Figure S5), similar to other reports in pre-clinical and clinical studies [55], [79], [80].

## 5. Supplemental Material

**Figure S1.**
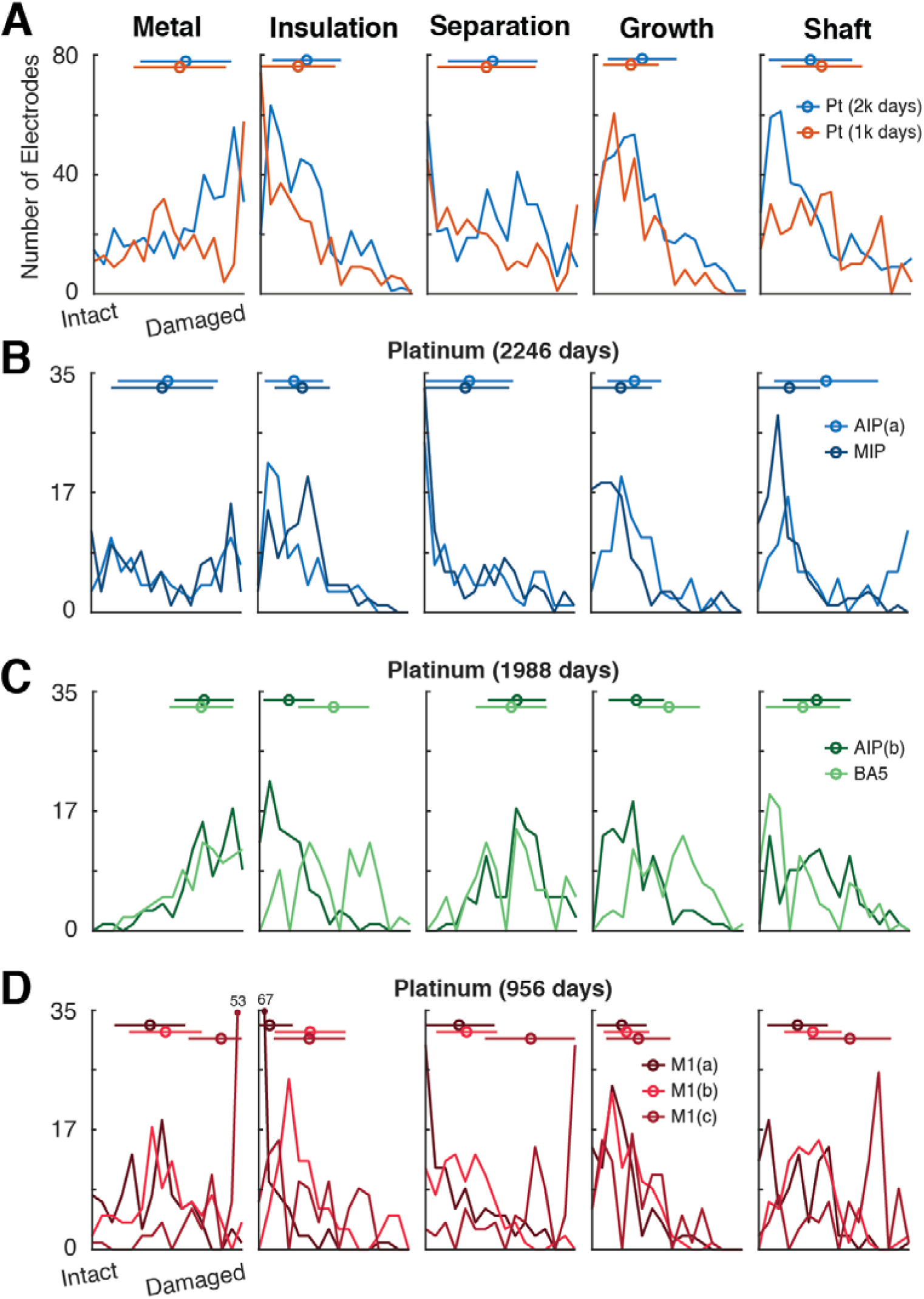
Detailed histogram of degradation metrics (Platinum) Detailed histogram of visually assessed degradation metrics from Figure 4A, separated by each individual array and participant. Mean and standard deviation are shown above as a line and a circle; however, many histograms appear to be bimodal or exponentially distributed. Y-axis shows the number of electrodes and X-axis shows level of damage (1, intact – 4, heavy damage). **(A)** Summary histogram of all Pt electrodes at ∼2000 days (blue) vs. ∼1000 days (red). Total number of electrodes at ∼2000 days was 389, and total number at ∼1000 days was 393. **(B-C)** Arrays from two participants at Caltech. **(D)** Pt arrays from one participant at JHU.

**Figure S2.**
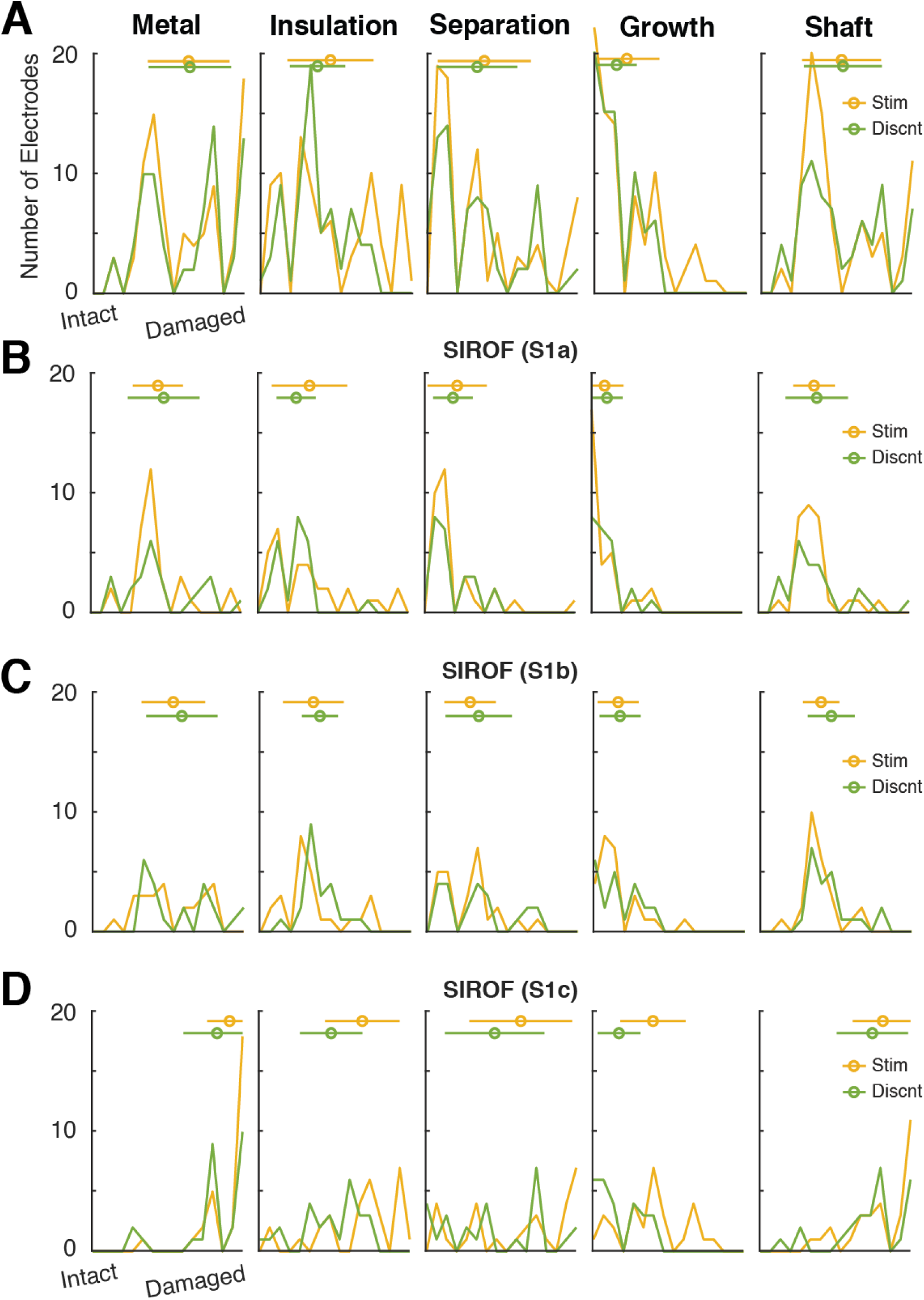
Detailed histogram of degradation metrics (SIROF) Detailed histogram of visually assessed degradation metrics from Figure 4B, separated by each individual array all from the JHU participant. Y-axis shows the number of electrodes and X-axis shows level of damage (1, intact – 4, heavy damage). **(A)** Summary histogram of all SIROF electrodes which were stimulated (yellow) vs. disconnected (green). See Figure 1F,G for spatial organization of electrodes. **(B-D)** Arrays from contralateral hemisphere. **(D)** One array from the ipsilateral hemisphere.

**Figure S3.**
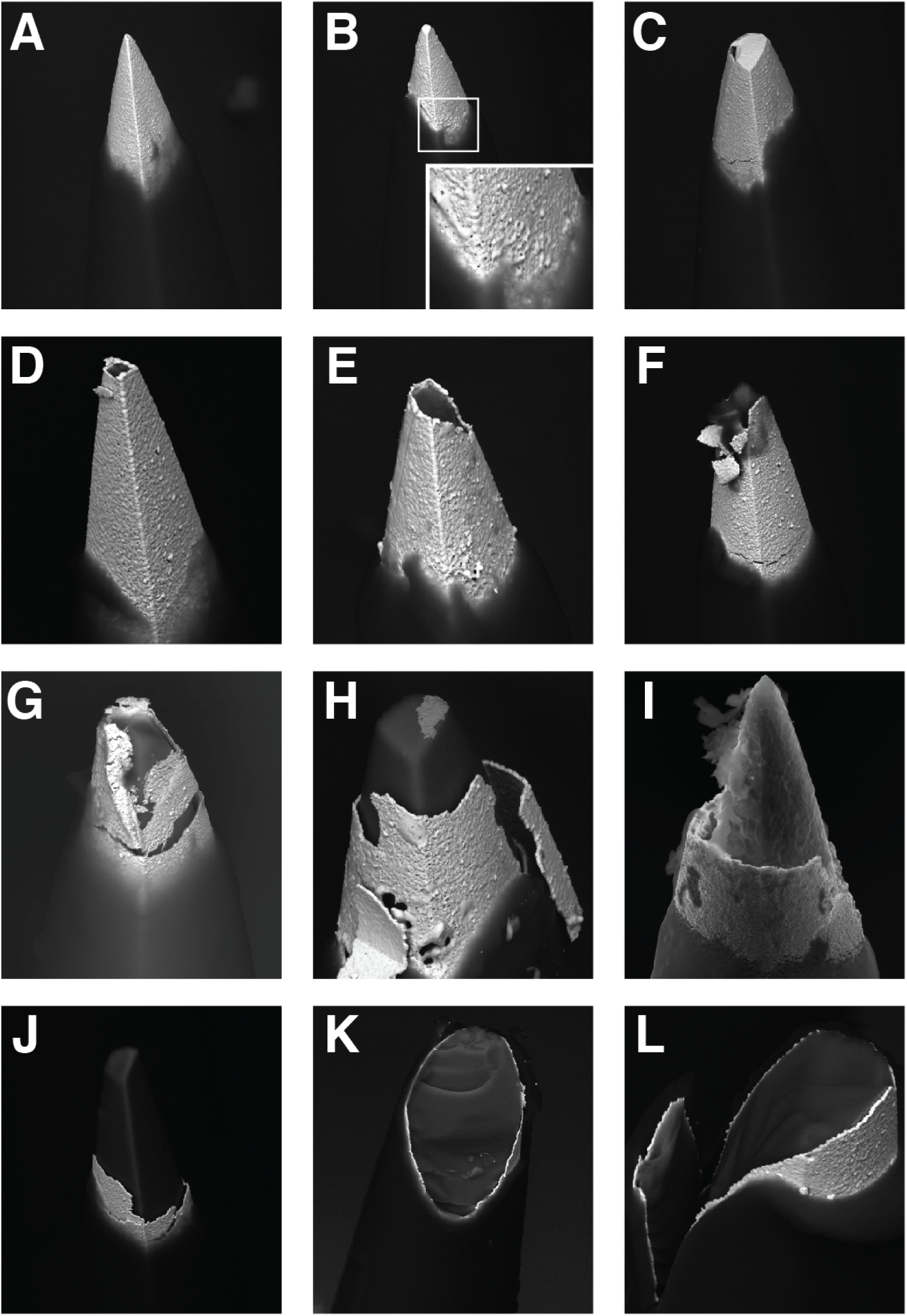
Example failure mechanisms, SEM images. We observed a wide variety of electrode metallization failure modes. Here are some examples of different states of degradation of the tip metal of these electrodes. **(A-C)** These electrodes were rated 1 (intact) for the metallization SEM assessment metric. They exhibited perfect or nearly perfect tip metal. Slight holes (inset in Panel B) or hairline fractures were allowed (Panel C). **(D-F)** These electrodes were rated 2 (slight damaged) and contained greater than 10% metal loss. In Panel F, the metal has begun to flake off the silicone shank. **(G-I)** These electrodes were rated 3 (damaged) and exhibited greater than 50% tip metal loss. The silicone beneath the metal has been etched away, reducing surface area and creating separation between the silicone and tip metal. **(J-L)** These electrodes were rated 4 (severely damaged). The metal is more than 90% lost. In Panel K,L, the silicone has also been complete eroded, leaving no support or surface area for the tip metal to adhere.

**Figure S4.**
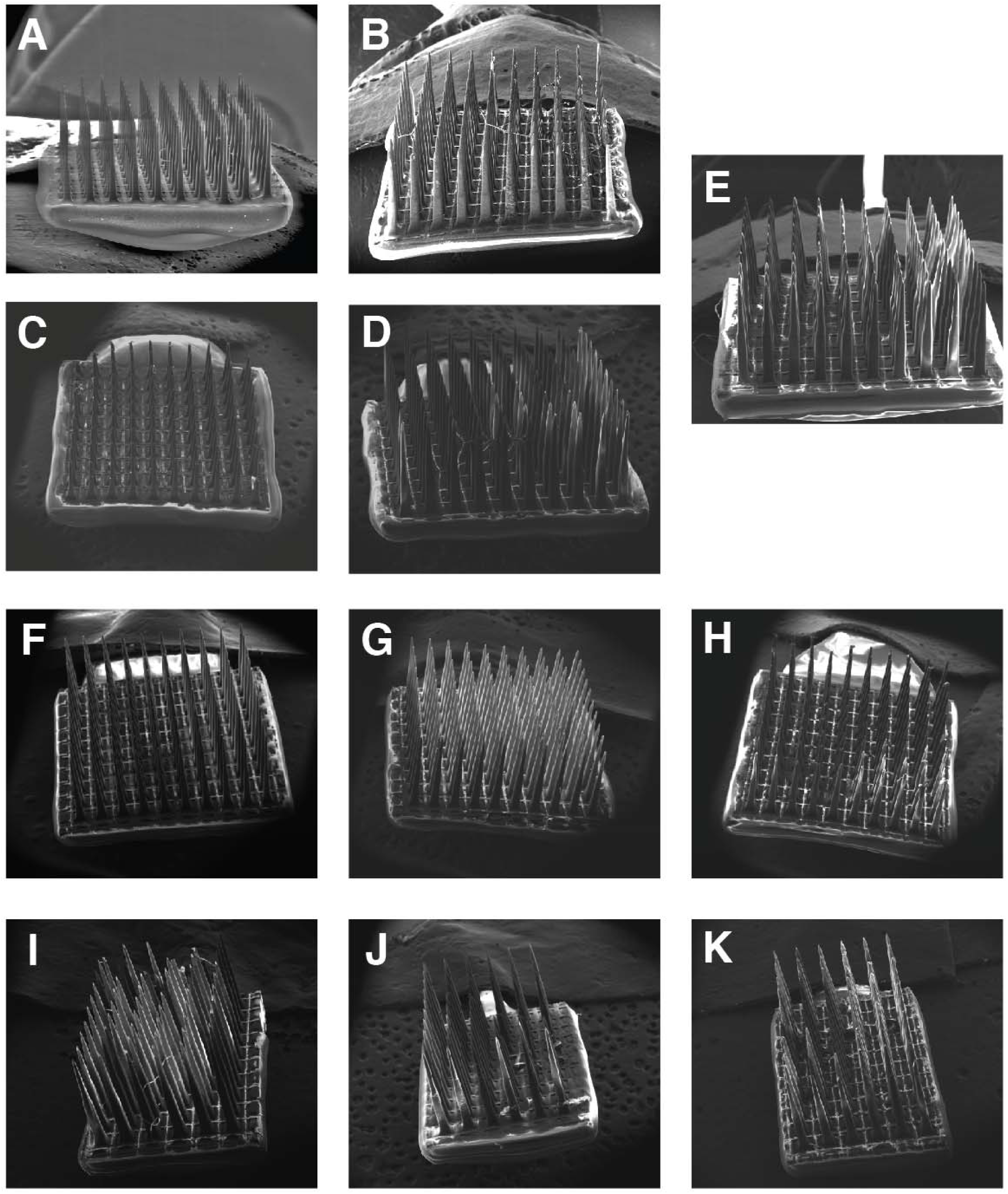
SEM images of each array. Each array was captured by SEM imaging. **(A)** AIP(b). **(B)** MIP. **(C)** AIP(a). **(D)** BA5. **(E)** Control. **(F)** M1(a). **(G)** M1(b). **(H)** M1(c). **(I)** S1(a). **(J)** S1(b). **(K)** S1(c).

**Figure S5.**
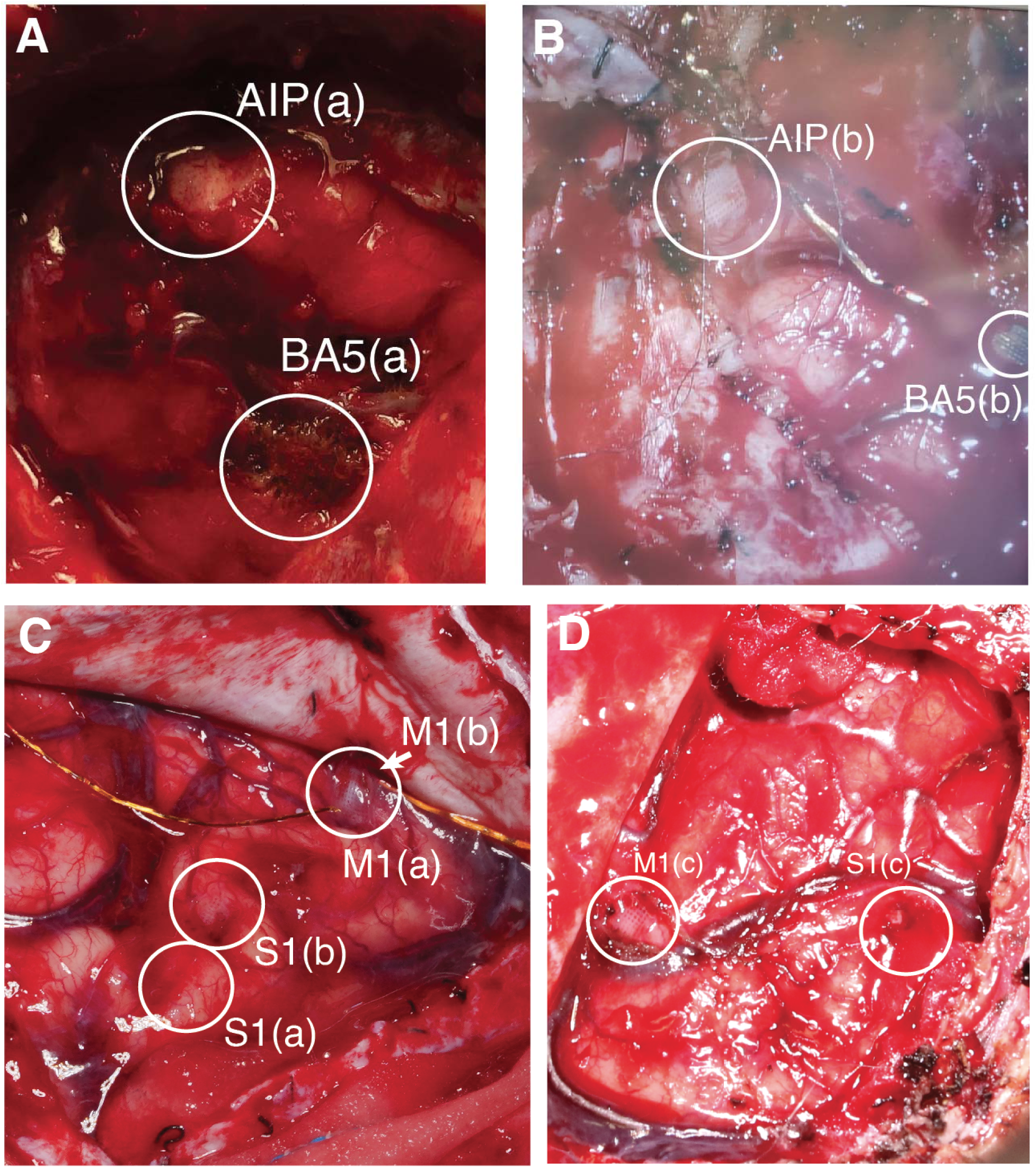
Post-explant cortex. Some imagery during the post explant procedures showed clear impressions of the shanks in the cortical tissue after the arrays were lifted out. **(A)** Caltech/UCLA **(B)** Caltech/USC **(C)** APL/JHU contralateral hemisphere. **(D)** APL/JHU ipsilateral hemisphere.

## Data Availability Statement

The data analyzed in this paper will be available upon request.

## Acknowledgements

The authors would like to thank all the participants for their efforts and engagement in the clinical study. We also thank the clinical staff at Rancho Los Amigos, Casa Colina, USC, UCLA and JHH their work and dedication during the experimental sessions. We would also like thank the staff at the University of Utah and Caltech Imaging Centers for their expertise and assistance imaging the arrays. Big thanks to BSS, JBV, BMG, KK and AS for their assistance scoring the SEM images. This work was developed with variety of funding support for salary, resources, patient care costs, and specialized equipment; including internal research support from the Johns Hopkins University Applied Physics Laboratory (JHU/APL) and funding from the Defense Advanced Research Projects Agency (DARPA) under awards HR001120C0120, N66001-10-C-4056 and N66001-15-C-4017. The views, opinions and/or findings expressed are those of the authors and should not be interpreted as representing the official views or policies of the Department of Defense or the U.S. Government.

## Funding

Tianqiao and Chrissy Chen Brain-machine Interface Center

Boswell Foundation

Craig H. Neilson Foundation

NIH/NINDS Grant U01NS098975

NIH/NINDS Grant U01NS123127

NIH/NEI R01EY015545

NIH/NEI UG1EY032039

NIH U01NS098975

DARPA N66001-10-C-4056

DARPA HR001120C0120

NIH/NEI: 1UG3NS107688

DARPA/BTO HAPTIX : N66001-15-C-4017

## Author Roles

Conceptualization: DB, SK, FT, LR, RAA

SEM Data Collection: DB, SK, LR, BB

End of Study Data Collection: DB, SK, RN, TA, LB, SC, MF, LO, BC, BW, PC, DK

Investigation: DB, SK, LR

Clinical: NC, WA, NP, BL, CL

Resources: KP, BB, FT, LR, RAA

Funding acquisition: DB, CL, FT, LR, RAA

Formal analysis: DB, SK

Writing - Original Draft: DB

Writing - Review & Editing: DB, SK, BC, FT, LR, RAA

## References

[1] E. M. Maynard, C. T. Nordhausen, and R. A. Normann, “The Utah Intracortical Electrode Array: A recording structure for potential brain-computer interfaces,” Electroencephalogr. Clin. Neurophysiol., vol. 102, no. 3, pp. 228–239, 1997, doi: 10.1016/S0013-4694(96)95176-0.

[2] P. R. Kennedy and R. A. E. Bakay, “Restoration of neural output from a paralyzed patient by a direct brain connection.,” Neuroreport, vol. 9, no. 8, pp. 1707–11, Jun. 1998, doi: 10.1097/00001756-199806010-00007.

[3] E. E. Fetz, “Real-time control of a robotic arm by neuronal ensembles,” Nat. Neurosci., vol. 2, no. 7, pp. 583–584, 1999, doi: 10.1038/10131.

[4] J. Wessberg et al., “Real-time prediction of hand tranjectory by ensembles of cortical neurons in primate,” Nature, vol. 408, no. 1, pp. 361–365, 2000.

[5] M. D. Serruya, N. G. Hatsopoulos, L. Paninski, M. R. Fellows, and J. P. Donoghue, “Instant neural control of a movement signal.,” Nature, vol. 416, no. 6877, pp. 141–2, Mar. 2002, doi: 10.1038/416141a.

[6] D. M. Taylor, “Direct Cortical Control of 3D Neuroprosthetic Devices,” Science (80-. )., vol. 296, no. 5574, pp. 1829–1832, Jun. 2002, doi: 10.1126/science.1070291.

[7] M. Velliste, S. Perel, M. C. Spalding, A. S. Whitford, and A. B. Schwartz, “Cortical control of a prosthetic arm for self-feeding,” Nature, vol. 453, no. 7198, pp. 1098–1101, 2008, doi: 10.1038/nature06996.

[8] L. R. Hochberg et al., “Reach and grasp by people with tetraplegia using a neurally controlled robotic arm.,” Nature, vol. 485, no. 7398, pp. 372–5, May 2012, doi: 10.1038/nature11076.

[9] J. L. Collinger et al., “High-performance neuroprosthetic control by an individual with tetraplegia,” Lancet, vol. 381, no. 9866, pp. 557–564, 2013, doi: 10.1016/S0140-6736(12)61816-9.

[10] B. Wodlinger, J. E. Downey, E. C. Tyler-Kabara, A. B. Schwartz, M. L. Boninger, and J. L. Collinger, “Ten-dimensional anthropomorphic arm control in a human brain−machine interface: difficulties, solutions, and limitations,” J. Neural Eng., vol. 12, no. 1, p. 016011, Feb. 2015, doi: 10.1088/1741-2560/12/1/016011.

[11] R. A. Andersen, T. Aflalo, L. Bashford, D. Bjånes, and S. Kellis, “Exploring Cognition with Brain–Machine Interfaces,” 10.1146/annurev-psych-030221-030214, vol. 73, no. 1, pp. 131–158, Jan. 2022, doi: 10.1146/ANNUREV-PSYCH-030221-030214.

[12] S. Musallam, B. D. Corneil, B. Greger, H. Scherberger, and R. A. Andersen, “Cognitive control signals for neural prosthetics,” Science (80-. )., vol. 305, no. 5681, pp. 258–262, 2004, doi: 10.1126/science.1097938.

[13] C. Pandarinath et al., “High performance communication by people with paralysis using an intracortical brain-computer interface,” Elife, vol. 6, Feb. 2017, doi: 10.7554/ELIFE.18554.

[14] S. K. Wandelt, D. A. Bjånes, K. Pejsa, B. Lee, C. Liu, and R. A. Andersen, “Online internal speech decoding from single neurons in a human participant,” medRxiv, p. 2022.11.02.22281775, Nov. 2022, doi: 10.1101/2022.11.02.22281775.

[15] R. A. Normann and E. Fernandez, “Clinical applications of penetrating neural interfaces and Utah Electrode Array technologies,” J. Neural Eng., vol. 13, no. 6, p. 061003, Oct. 2016, doi: 10.1088/1741-2560/13/6/061003.

[16] M. A. Salas et al., “Proprioceptive and cutaneous sensations in humans elicited by intracortical microstimulation,” Elife, vol. 7, 2018, doi: 10.7554/eLife.32904.

[17] S. N. Flesher et al., “Intracortical microstimulation of human somatosensory cortex,” Sci. Transl. Med., vol. 8, no. 361, p. 361ra141, Oct. 2016, doi: 10.1126/scitranslmed.aaf8083.

[18] M. S. Fifer et al., “Intracortical Somatosensory Stimulation to Elicit Fingertip Sensations in an Individual With Spinal Cord Injury,” Neurology, p. 10.1212/WNL.0000000000013173, 2021, doi: 10.1212/wnl.0000000000013173.

[19] E. Fernández et al., “Visual percepts evoked with an intracortical 96-channel microelectrode array inserted in human occipital cortex,” J. Clin. Invest., vol. 131, no. 23, Dec. 2021, doi: 10.1172/JCI151331.

[20] X. Chen, F. Wang, E. Fernandez, and P. R. Roelfsema, “Shape perception via a high-channel-count neuroprosthesis in monkey visual cortex,” Science (80-. )., vol. 370, no. 6521, Dec. 2020, doi: 10.1126/SCIENCE.ABD7435/SUPPL_FILE/ABD7435S6.MP4.

[21] C. L. Hughes et al., “Neural stimulation and recording performance in human sensorimotor cortex over 1500 days,” J. Neural Eng., vol. 18, no. 4, Aug. 2021, doi: 10.1088/1741-2552/AC18AD.

[22] J. D. Simeral, S. P. Kim, M. J. Black, J. P. Donoghue, and L. R. Hochberg, “Neural control of cursor trajectory and click by a human with tetraplegia 1000 days after implant of an intracortical microelectrode array,” in Journal of Neural Engineering, Apr. 2011, vol. 8, no. 2. doi: 10.1088/1741-2560/8/2/025027.

[23] J. E. Downey, N. Schwed, S. M. Chase, A. B. Schwartz, and J. L. Collinger, “Intracortical recording stability in human brain-computer interface users,” J. Neural Eng., vol. 15, no. 4, May 2018, doi: 10.1088/1741-2552/AAB7A0.

[24] X. Chen et al., “Chronic stability of a neuroprosthesis comprising multiple adjacent Utah arrays in monkeys,” J. Neural Eng., vol. 20, no. 3, Jun. 2023, doi: 10.1088/1741-2552/ACE07E.

[25] S. Suner, M. R. Fellows, C. Vargas-Irwin, G. K. Nakata, and J. P. Donoghue, “Reliability of signals from a chronically implanted, silicon-based electrode array in non-human primate primary motor cortex,” IEEE Trans. Neural Syst. Rehabil. Eng., vol. 13, no. 4, pp. 524–541, 2005, doi: 10.1109/TNSRE.2005.857687.

[26] C. A. Chestek et al., “Long-term stability of neural prosthetic control signals from silicon cortical arrays in rhesus macaque motor cortex,” J. Neural Eng., vol. 8, no. 4, Aug. 2011, doi: 10.1088/1741-2560/8/4/045005.

[27] J. D. Simeral, S. P. Kim, M. J. Black, J. P. Donoghue, and L. R. Hochberg, “Neural control of cursor trajectory and click by a human with tetraplegia 1000 days after implant of an intracortical microelectrode array,” J. Neural Eng., vol. 8, no. 2, 2011, doi: 10.1088/1741-2560/8/2/025027.

[28] J. C. Barrese et al., “Failure mode analysis of silicon-based intracortical microelectrode arrays in non-human primates,” J. Neural Eng., vol. 10, no. 6, 2013, doi: 10.1088/1741-2560/10/6/066014.

[29] J. A. George et al., “Long-term performance of Utah slanted electrode arrays and intramuscular electromyographic leads implanted chronically in human arm nerves and muscles,” J. Neural Eng., vol. 17, no. 5, p. 056042, Oct. 2020, doi: 10.1088/1741-2552/ABC025.

[30] M. M. Straka, B. Shafer, S. Vasudevan, C. Welle, and L. Rieth, “Characterizing Longitudinal Changes in the Impedance Spectra of In-Vivo Peripheral Nerve Electrodes,” Micromachines 2018, Vol. 9, Page 587, vol. 9, no. 11, p. 587, Nov. 2018, doi: 10.3390/MI9110587.

[31] A. Prasad et al., “Abiotic-biotic characterization of Pt/Ir microelectrode arrays in chronic implants,” Front. Neuroeng., vol. 7, no. FEB, p. 2, Feb. 2014, doi: 10.3389/FNENG.2014.00002/BIBTEX.

[32] T. D. Y. Kozai et al., “Mechanical failure modes of chronically implanted planar silicon-based neural probes for laminar recording,” Biomaterials, vol. 37, pp. 25–39, Jan. 2015, doi: 10.1016/J.BIOMATERIALS.2014.10.040.

[33] K. Woeppel, Q. Yang, and X. T. Cui, “Recent advances in neural electrode–tissue interfaces,” Curr. Opin. Biomed. Eng., vol. 4, pp. 21–31, Dec. 2017, doi: 10.1016/J.COBME.2017.09.003.

[34] V. S. Polikov, P. A. Tresco, and W. M. Reichert, “Response of brain tissue to chronically implanted neural electrodes,” J. Neurosci. Methods, vol. 148, no. 1, pp. 1–18, 2005, doi: 10.1016/j.jneumeth.2005.08.015.

[35] J. W. Salatino, K. A. Ludwig, T. D. Y. Kozai, and E. K. Purcell, “Glial responses to implanted electrodes in the brain,” Nat. Biomed. Eng., vol. 1, no. 11, pp. 862–877, Nov. 2017, doi: 10.1038/S41551-017-0154-1.

[36] L. J. Szymanski et al., “Neuropathological effects of chronically implanted, intracortical microelectrodes in a tetraplegic patient,” J. Neural Eng., vol. 18, no. 4, pp. 460–469, Aug. 2021, doi: 10.1088/1741-2552/ac127e.

[37] A. Woolley, H. Desai, K. O.-J. of neural engineering, and undefined 2013, “Chronic intracortical microelectrode arrays induce non-uniform, depth-related tissue responses,” iopscience.iop.orgAJ Woolley, HA Desai, KJ OttoJournal neural Eng. 2013•iopscience.iop.org, vol. 10, no. 2, Apr. 2013, doi: 10.1088/1741-2560/10/2/026007.

[38] J. C. Barrese, J. Aceros, and J. P. Donoghue, “Scanning electron microscopy of chronically implanted intracortical microelectrode arrays in non-human primates,” J. Neural Eng., vol. 13, no. 2, p. 026003, Jan. 2016, doi: 10.1088/1741-2560/13/2/026003.

[39] C. F. Dunlap, S. C. Colachis, E. C. Meyers, M. A. Bockbrader, and D. A. Friedenberg, “Classifying Intracortical Brain-Machine Interface Signal Disruptions Based on System Performance and Applicable Compensatory Strategies: A Review,” Front. Neurorobot., vol. 14, Oct. 2020, doi: 10.3389/FNBOT.2020.558987/FULL.

[40] G. Buzsáki, “Large-scale recording of neuronal ensembles,” Nat. Neurosci., vol. 7, no. 5, pp. 446–451, May 2004, doi: 10.1038/NN1233.

[41] A. B. Schwartz, X. T. Cui, D. J. J. Weber, and D. W. Moran, “Brain-Controlled Interfaces: Movement Restoration with Neural Prosthetics,” Neuron, vol. 52, no. 1, pp. 205–220, 2006, doi: 10.1016/j.neuron.2006.09.019.

[42] P. J. Rousche and R. A. Normann, “Chronic recording capability of the utah intracortical electrode array in cat sensory cortex,” J. Neurosci. Methods, vol. 82, no. 1, pp. 1–15, 1998, doi: 10.1016/S0165-0270(98)00031-4.

[43] A. Degenhart, J. Eles, R. Dum, … J. M.-J. of neural, and undefined 2016, “Histological evaluation of a chronically-implanted electrocorticographic electrode grid in a non-human primate,” iopscience.iop.orgAD Degenhart, J Eles, R Dum, JL Mischel, I Smalianchuk, B Endler, RC AshmoreJournal neural Eng. 2016•iopscience.iop.org, vol. 13, no. 4, Jun. 2016, doi: 10.1088/1741-2560/13/4/046019.

[44] E. M. Schmidt, J. S. Mcintosh, and M. J. Bak, “Long-term implants of Parylene-C coated microelectrodes,” Med. Biol. Eng. Comput., vol. 26, no. 1, pp. 96–101, Jan. 1988, doi: 10.1007/BF02441836.

[45] X. Xie et al., “Long-term reliability of Al2O3 and Parylene C bilayer encapsulated Utah electrode array based neural interfaces for chronic implantation,” J. Neural Eng., vol. 11, no. 2, p. 9, 2014, doi: 10.1088/1741-2560/11/2/026016.

[46] R. Caldwell, M. G. Street, R. Sharma, P. Takmakov, B. Baker, and L. Rieth, “Characterization of Parylene-C degradation mechanisms: In vitro reactive accelerated aging model compared to multiyear in vivo implantation,” Biomaterials, vol. 232, p. 119731, Feb. 2020, doi: 10.1016/J.BIOMATERIALS.2019.119731.

[47] R. Caldwell et al., “Neural electrode resilience against dielectric damage may be improved by use of highly doped silicon as a conductive material,” J. Neurosci. Methods, vol. 293, pp. 210–225, Jan. 2018, doi: 10.1016/J.JNEUMETH.2017.10.002.

[48] P. Ghelich, N. F. Nolta, and M. Han, “Unprotected sidewalls of implantable silicon-based neural probes and conformal coating as a solution,” Npj Mater. Degrad., vol. 5, no. 1, Dec. 2021, doi: 10.1038/S41529-021-00154-9.

[49] S. Negi, R. Bhandari, L. Rieth, and F. Solzbacher, “In vitro comparison of sputtered iridium oxide and platinum-coated neural implantable microelectrode arrays.,” Biomed. Mater., vol. 5, no. 1, p. 15007, 2010, doi: 10.1088/1748-6041/5/1/015007.

[50] S. Negi, R. Bhandari, R. Van Wagenen, and F. Solzbacher, “Factors affecting degradation of sputtered iridium oxide used for neuroprosthetic applications,” Proc. IEEE Int. Conf. Micro Electro Mech. Syst., no. c, pp. 568–571, 2010, doi: 10.1109/MEMSYS.2010.5442438.

[51] A. Ghazavi and S. F. Cogan, “Ultramicro-sized sputtered iridium oxide electrodes in buffered saline: Behavior, stability, and the effect of the perimeter to area ratio on their electrochemical properties,” Electrochim. Acta, vol. 423, p. 140514, Aug. 2022, doi: 10.1016/J.ELECTACTA.2022.140514.

[52] T. Sun et al., “Flexible IrOx neural electrode for mouse vagus nerve stimulation,” Acta Biomater., vol. 159, pp. 394–409, Mar. 2023, doi: 10.1016/J.ACTBIO.2023.01.026.

[53] S. F. Cogan, “Neural Stimulation and Recording Electrodes,” Annu. Rev. Biomed. Eng., vol. 10, no. 1, pp. 275–309, 2008, doi: 10.1146/annurev.bioeng.10.061807.160518.

[54] P. J. Gilgunn, X. C. Ong, S. N. Flesher, A. B. Schwartz, and R. A. Gaunt, “Structural analysis of explanted microelectrode arrays,” Int. IEEE/EMBS Conf. Neural Eng. NER, pp. 719–722, 2013, doi: 10.1109/NER.2013.6696035.

[55] K. Woeppel et al., “Explant Analysis of Utah Electrode Arrays Implanted in Human Cortex for Brain-Computer-Interfaces,” Front. Bioeng. Biotechnol., vol. 9, p. 1137, Dec. 2021, doi: 10.3389/FBIOE.2021.759711/BIBTEX.

[56] P. Takmakov, K. Ruda, K. Scott Phillips, I. S. Isayeva, V. Krauthamer, and C. G. Welle, “Rapid evaluation of the durability of cortical neural implants using accelerated aging with reactive oxygen species,” J. Neural Eng., vol. 12, no. 2, Apr. 2015, doi: 10.1088/1741-2560/12/2/026003.

[57] M. G. Street, C. G. Welle, and P. A. Takmakov, “Automated reactive accelerated aging for rapid in vitro evaluation of neural implant performance,” Rev. Sci. Instrum., vol. 89, no. 9, Sep. 2018, doi: 10.1063/1.5024686.

[58] A. Prasad et al., “Comprehensive characterization and failure modes of tungsten microwire arrays in chronic neural implants,” J. Neural Eng., vol. 9, no. 5, Oct. 2012, doi: 10.1088/1741-2560/9/5/056015.

[59] A. Prasad and J. C. Sanchez, “Quantifying long-term microelectrode array functionality using chronic in vivo impedance testing,” J. Neural Eng., vol. 9, no. 2, Apr. 2012, doi: 10.1088/1741-2560/9/2/026028.

[60] C. Bennett, M. Samikkannu, F. Mohammed, W. D. Dietrich, S. M. Rajguru, and A. Prasad, “Blood brain barrier (BBB)-disruption in intracortical silicon microelectrode implants,” Biomaterials, vol. 164, pp. 1–10, May 2018, doi: 10.1016/j.biomaterials.2018.02.036.

[61] C. Bennett et al., “Neuroinflammation, oxidative stress, and blood-brain barrier (BBB) disruption in acute Utah electrode array implants and the effect of deferoxamine as an iron chelator on acute foreign body response,” Biomaterials, vol. 188, pp. 144–159, Jan. 2019, doi: 10.1016/J.BIOMATERIALS.2018.09.040.

[62] S. Negi, R. Bhandari, L. Rieth, and F. Solzbacher, “In vitro comparison of sputtered iridium oxide and platinum-coated neural implantable microelectrode arrays.,” Biomed. Mater., vol. 5, no. 1, p. 15007, 2010, doi: 10.1088/1748-6041/5/1/015007.

[63] T. Aflalo et al., “Neurophysiology. Decoding motor imagery from the posterior parietal cortex of a tetraplegic human.,” Science, vol. 348, no. 6237, pp. 906–10, May 2015, doi: 10.1126/science.aaa5417.

[64] T. Aflalo, C. Zhang, B. Revechkis, E. Rosario, N. Pouratian, and R. A. Andersen, “Implicit mechanisms of intention,” Curr. Biol., vol. 32, no. 9, pp. 2051–2060.e6, May 2022, doi: 10.1016/J.CUB.2022.03.047.

[65] A. T. Gardner, H. J. Strathman, D. J. Warren, and R. M. Walker, “Impedance and noise characterizations of Utah and microwire electrode arrays,” IEEE J. Electromagn. RF Microwaves Med. Biol., vol. 2, no. 4, pp. 234–241, Dec. 2018, doi: 10.1109/JERM.2018.2862417.

[66] T. Chai and R. R. Draxler, “Root mean square error (RMSE) or mean absolute error (MAE)?-Arguments against avoiding RMSE in the literature,” Geosci. Model Dev, vol. 7, pp. 1247–1250, 2014, doi: 10.5194/gmd-7-1247-2014.

[67] P. A. P. Moran, “Notes on Continuous Stochastic Phenomena,” Biometrika, vol. 37, no. 1/2, p. 17, Jun. 1950, doi: 10.2307/2332142.

[68] S. F. Cogan, “Neural Stimulation and Recording Electrodes,” Annu. Rev. Biomed. Eng., vol. 10, no. 1, pp. 275–309, Aug. 2008, doi: 10.1146/annurev.bioeng.10.061807.160518.

[69] A. T. Rajan et al., “The effects of chronic intracortical microstimulation on neural tissue and fine motor behavior,” J. Neural Eng., vol. 12, no. 6, p. 66018, 2015, doi: 10.1088/1741-2560/12/6/066018.

[70] K. A. Sillay et al., “Long-Term Measurement of Impedance in Chronically Implanted Depth and Subdural Electrodes During Responsive Neurostimulation in Humans,” Brain Stimul., vol. 6, no. 5, pp. 718–726, Sep. 2013, doi: 10.1016/J.BRS.2013.02.001.

[71] S. A. Jones, S.-H. Shim, J. He, and X. Zhuang, “Fast, three-dimensional super-resolution imaging of live cells,” Nat. Methods, vol. 8, no. 6, pp. 499–505, 2011, doi: 10.1038/nmeth.1605.

[72] V. Thakore, P. Molnar, and J. J. Hickman, “An optimization-based study of equivalent circuit models for representing recordings at the neuron-electrode interface,” IEEE Trans. Biomed. Eng., vol. 59, no. 8, pp. 2338–2347, 2012, doi: 10.1109/TBME.2012.2203820.

[73] N. Lago et al., “A physical-based equivalent circuit model for an organic/electrolyte interface,” Org. Electron., vol. 35, pp. 176–185, Aug. 2016, doi: 10.1016/J.ORGEL.2016.05.018.

[74] J. Jiang, F. R. Willett, and D. M. Taylor, “Relationship between microelectrode array impedance and chronic recording quality of single units and local field potentials,” 2014 36th Annu. Int. Conf. IEEE Eng. Med. Biol. Soc. EMBC 2014, pp. 3045–3048, Nov. 2014, doi: 10.1109/EMBC.2014.6944265.

[75] P. A. Cody, J. R. Eles, C. F. Lagenaur, T. D. Y. Kozai, and X. T. Cui, “Unique electrophysiological and impedance signatures between encapsulation types: An analysis of biological Utah array failure and benefit of a biomimetic coating in a rat model,” Biomaterials, vol. 161, pp. 117–128, Apr. 2018, doi: 10.1016/J.BIOMATERIALS.2018.01.025.

[76] S. W. . Freiman and J. J. . Mecholsky, The Fracture of Brittle Materials: Testing and Analysis. John Wiley & Sons, Inc., 2012. doi: 10.1002/9781118147757.

[77] E. Ilic, A. Pardo, R. Hauert, P. Schmutz, and S. Mischler, “Silicon Corrosion in Neutral Media: The Influence of Confined Geometries and Crevice Corrosion in Simulated Physiological Solutions,” J. Electrochem. Soc., vol. 166, no. 6, pp. C125–C133, Mar. 2019, doi: 10.1149/2.0241906JES/XML.

[78] S. R. Kane, S. F. Cogan, J. Ehrlich, T. D. Plante, D. B. McCreery, and P. R. Troyk, “Electrical performance of penetrating microelectrodes chronically implanted in cat cortex,” IEEE Trans. Biomed. Eng., vol. 60, no. 8, pp. 2153–2160, 2013, doi: 10.1109/TBME.2013.2248152.

[79] P. A. House, J. D. MacDonald, P. A. Tresco, and R. A. Normann, “Acute microelectrode array implantation into human neocortex: preliminary technique and histological considerations,” Neurosurg. Focus, vol. 20, no. 5, 2006, doi: 10.3171/FOC.2006.20.5.5.

[80] B. P. Christie, K. R. Ashmont, P. A. House, and B. Greger, “Approaches to a cortical vision prosthesis: implications of electrode size and placement,” J. Neural Eng., vol. 13, no. 2, Feb. 2016, doi: 10.1088/1741-2560/13/2/025003.

